# Single-cell multi-omic profiling allows the dissection of peripheral immune phenotypes in Alzheimer’s Disease progression

**DOI:** 10.64898/2026.03.12.26348228

**Authors:** Jannis B Spintge, Karola Mai, Caterina Carraro, Martina van Uelft, Francesco Elli, Karoline Mauer, Lisa Holsten, Aleksej Frolov, Janice Elangikal, Emily Hinkley, Jonas Schulte-Schrepping, Mehrnoush H Shakiba, Laura Lang, Tarek Elmzzahi, Doaa Hamada, Sophie Müller, Yuanfang Li, Ioanna Gemünd, Charlotte Kröger, Jacqueline Leidner, Timo Zajac, Jessica V Montgomery, Celia Hartmann, Bidour Hussein, Maren Büttner, Rainer Knoll, Dina Hüsson, Rebekka Scholz, Tobias Paschek, Victoria Isakzai, Nico Reusch, Stefan Paulusch, Anna Drews, Michael Kraut, Heidi Theis, Marie Rüthing, Ulrike Strube, Lukas Preis, Daria Gref, Eike J Spruth, Maria Gemenetzi, Klaus Fliessbach, Niels Hansen, Ayda Rostamzadeh, Wenzel Glanz, Enise I Incesoy, Daniel Janowitz, Sophia Stöcklein, Boris-Stephan Rauchmann, Ingo Kilimann, Doreen Goerss, Sebastian Sodenkamp, Marie Kronmüller, Sandra Roeske, Anna Gamez, Matthias Schmid, Katharina Bürger, Julian Hellmann-Regen, Christoph Laske, Alfredo Ramirez, Frederic Brosseron, Jens Wiltfang, Robert Perneczky, Emrah Düzel, Stefan Teipel, Annika Spottke, Gabor C Petzold, Oliver Peters, Josef Priller, Susanne V Schmidt, Michael Wagner, Luca Kleineidam, Frank Jessen, Anja Schneider, Lorenzo Bonaguro, Elena De Domenico, Anna C Aschenbrenner, Matthias Becker, Thomas Ulas, Joachim L Schultze, Marc D Beyer

## Abstract

The role of the peripheral immune system in Alzheimer’s Disease (AD) remains insufficiently resolved, limiting the understanding of systemic disease effects and mechanisms. Here, we employed three high-resolution single-cell techniques, including flow cytometry, single-cell RNA- and ATAC-sequencing, to investigate peripheral immunity in AD dementia and earlier stages of the AD trajectory in over 100 patients. We identified reduced humoral immune responses in AD, characterized by a diminished B cell compartment displaying an impaired activation phenotype. Classical monocytes expanded in mild cognitive impairment and early AD dementia, acquiring a NF-*k*B/AP-1-mediated low-grade inflammation phenotype. Our findings link peripheral dysregulation in innate and adaptive immunity at cell frequency, transcriptional and epigenetic levels to the AD trajectory and provide insights into distinct phenotypes that define AD progression in contrast to healthy aging across cohorts.

## Introduction

Alzheimer’s disease (AD) is a progressive neurodegenerative disorder and the leading cause of dementia worldwide, accounting for 60-70% of all cases^1^. Over 50 million people are affected globally, and with aging populations driving projections to triple this number by 2050, AD imposes a significant personal, economic, and healthcare burden with annual global costs exceeding $1 trillion^2,3^. Clinically, AD is characterized by progressive cognitive decline involving memory loss, impaired reasoning, and behavioural changes^1^. Pathologically, the disease is defined by hallmark features including amyloid-beta (Aβ) plaque accumulation and neurofibrillary tangles composed of hyperphosphorylated tau protein^4^ in the brain. These pathological events act synergistically with neuroinflammatory processes and contribute to synaptic dysfunction and neuronal loss^5^. While aging remains the primary risk factor, genetic predisposition also contributes to AD risk^6^. Moreover, certain environmental factors such as microbiome, physical activity, and systemic inflammation influence disease onset and progression^7–10^. Advances in biomarker discovery have enhanced early detection capabilities through cerebrospinal fluid (CSF) analyses that can detect Aβ peptides and tau proteins up to a decade before the appearance of clinical symptoms^11^.

Growing evidence suggests that immune dysregulation is not merely a consequence but an active driver of AD pathogenesis^12,13^. Dysfunction in both innate and adaptive immunity contributes to neuroinflammation and misfolded Aβ and tau proteins can trigger immune responses that exacerbate inflammation^14^. Genome-wide association studies (GWAS) have identified immune-related loci that increase AD risk by affecting immune cell function^15,16^. Potential interactions with peripheral immunity have also been described^14,17,18^, such as AD-associated changes in peripheral adaptive immune cells^19–21^. While the infiltration of the central nervous system (CNS) by specific terminally differentiated and expanded CD8+ T cell subsets has been established for AD^14^, other peripheral immune compartments lack a known consensus mechanism of action in the disease context. In particular, peripheral monocytes have been suggested to play a role in both exacerbating neuroinflammation and ameliorating Aβ pathology, giving them an ambiguous role in AD^22–25^. Together, exact mechanisms how peripheral immunity impacts AD pathogenesis are still under debate, and conflicting roles for multiple immune cell types have been reported^18^ but only a limited number of studies have focused on peripheral blood using high-dimensional and high-resolution multi-omics approaches^26–28^. To address this gap and to capture the full spectrum of the AD trajectory which ranges from subjective cognitive decline (SCD) through the prodromal stage of mild cognitive impairment (MCI) to established AD dementia, the German Center for Neurodegenerative Diseases (DZNE) initiated a multi-center longitudinal study (DELCODE). Within this framework, a sub-cohort comprising peripheral blood samples from Aβ-positive individuals with AD and varying degrees of cognitive impairment, as well as Aβ-negative controls without cognitive impairment were defined for the CureDem study. By employing multi-omic single-cell analysis including cell state proportions, transcriptomes, and epigenomes, we delineated cellular phenotypes across B cell, T cell and monocyte compartments associated with AD trajectories across donors. Our work provides novel insights into the complex interplay between peripheral immunity and neurodegeneration in AD and sets the stage for future investigations aimed at therapeutic targeting of immune pathways.

## Results

### Multi-omics reveal immune modulation across peripheral immune compartments associated with different stages of the Alzheimer’s Disease trajectory

To investigate the interplay of AD and peripheral immunity, we performed a multi-omic characterization of peripheral blood mononuclear cells (PBMCs) from 81 individuals recruited within the DELCODE/DESCRIBE studies of the DZNE (Figure 1A-B, Table S1). Cognitive staging of participants was primarily based on extensive neuropsychological testing (see methods), participants were further staged using the amyloid-tau-neurodegeneration (ATN) framework^29^, and stratification was based on CSF Aβ status^30^ (Table S1). The discovery cohort comprised 33 AD dementia (ADD) patients, 13 individuals with MCI due to AD, 11 subjects with SCD due to AD, all of whom were Aβ-positive (A+), along with 24 cognitively healthy controls who were Aβ-negative (A-) (Figure 1A-B, Table S1). The majority of individuals in the ADD and MCI groups carried at least one ApoE4 risk allele and presented with higher cognitive impairment compared to the SCD and control groups (Figure 1B, Table S1). After thorough modality-wise quality control, 66 samples were assessed by multi-color flow cytometry (MCFC), 69 samples by single-cell RNA sequencing (scRNA-seq) and 71 samples by single-cell assay for transposase-accessible chromatin using sequencing (scATAC-seq), respectively (Figure 1A). We obtained 24.7 million cells for MCFC (downsampled to 397,768) (Figure 1C), 185,246 cells for scRNA-seq (Figure 1D) and 76,061 single nuclei for scATAC-seq (Figure 1E).

**Figure 1 –.**
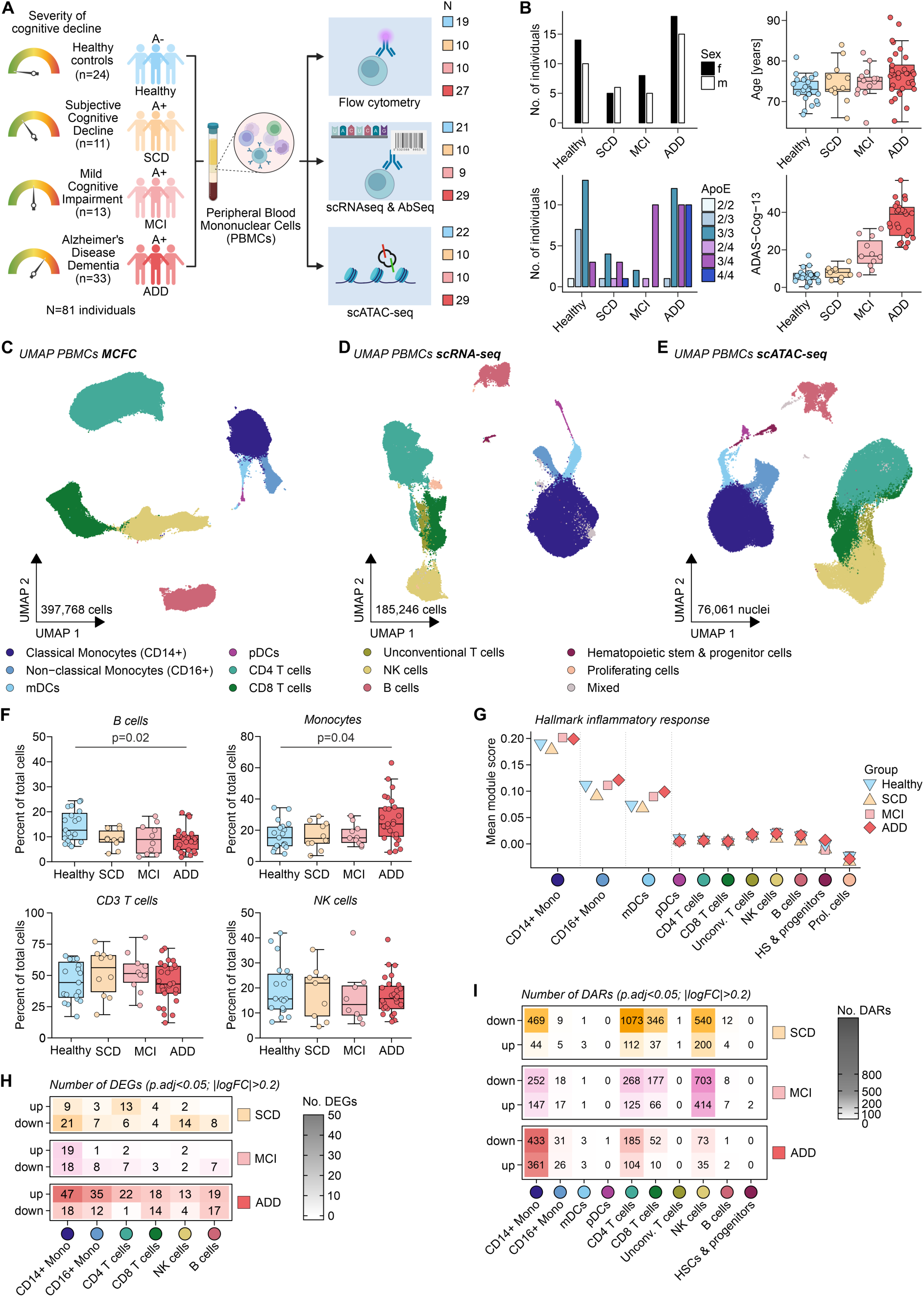
Multi-omics reveal immune modulation across peripheral immune compartments associated with different stages of the Alzheimer’s Disease trajectory. (A) Overview of the CureDem study design. Amyloid-positive patients with different stages of Alzheimer’s Disease with varying cognitive decline ranging from subjective cognitive decline (SCD) to mild cognitive impairment (MCI) up to mild Alzheimer’s Disease dementia (ADD) as well as cognitively healthy amyloid-negative controls underwent peripheral blood sampling. Peripheral blood mononuclear cells (PBMCs) were analyzed using multi-color flow cytometry (MCFC), scRNA-seq paired with AbSeq, and scATAC-seq. Created with BioRender. (B) Overview of study cohort demographics, in particular distributions of sex, age, ApoE alleles and Alzheimer’s Disease Assessment Scale cognitive subscale 13 (ADAS-Cog-13) scores per group. (C) UMAP visualization of the MCFC dataset (n = 24,684,219 cells were downsampled to 397,768 cells for visualization) paired scRNA-seq/AbSeq dataset (n = 185,246 cells) (D) and scATAC-seq dataset (n = 76,061 nuclei) (E), colored by concordant main cell type annotation. (F) Relative frequencies of annotated main cell types per group in the MCFC dataset. Percentages are relative to all CD45+ live cells. Statistical analysis using Kruskal-Wallis and post hoc Dunn’s test, Benjamini-Hochberg-adjusted p-values are indicated. (G) Module scores of the enrichment of the hallmark database *inflammatory response* gene set per group and main cell type. (H) Number of differentially expressed genes (DEGs) compared to controls per main cell type (with > 5,000 total cells) and group. Differential expression defined by Bonferroni-adjusted p-value < 0.05 and absolute average log2 fold change > 0.2 (iterative logistic regression framework with correction for season of sampling and sample of origin). (I) Number of differentially accessible regions (DARs) compared to controls per main cell type and group. Differential accessibility defined by Bonferroni-adjusted p-value < 0.05 and absolute average log2 fold change > 0.2.

All major PBMC subsets across all modalities were defined by unsupervised clustering and subsequent annotation guided by literature-validated cell type-defining markers (Figures 1C-E, S1A-D). MCFC profiling of cell type composition revealed a significant reduction in B cell frequencies among ADD patients relative to controls and a simultaneous expansion of the monocyte compartment, while other immune compartments were largely unaffected (Figure 1F). The loss of B cells in the ADD group compared to controls was consistently recapitulated in both the scRNA-seq and scATAC-seq datasets (Figure S1E). Overall, the observed cell type distributions did not show statistically significant differences concerning age, ApoE genotype and sex, only the increase in monocytes was more prominent in female ADD donors (Figure S1F-H).

Differential gene expression analysis and gene set enrichment analysis (GSEA) further demonstrated that classical monocytes exhibited the highest extent of overall gene expression modulation and the most robust expression of an inflammatory signature across all identified immune cell types (Figure 1G-H). For ADD and MCI patients, we observed that the average inflammatory profile of classical monocytes, and to a lesser extent non-classical monocytes and dendritic cells (DCs), was elevated, positioning the myeloid compartment as a key driver of inflammation within the AD trajectory (SCD>MCI>ADD) (Figure 1G, *square+diamond*). Similar to cell type distributions, gene expression patterns were not majorly influenced by demographic factors (Figure S1I).

As epigenetic changes were also previously suggested as potential contributors to functional reprogramming of cells in AD^31^, we assessed epigenetic profiles by examining differential chromatin accessibility across cell types. This analysis confirmed the most prominent remodelling in classical monocytes for individuals with ADD (Figure 1I), as well as dysregulation within the lymphoid compartment along the AD trajectory, in line with previous reports^28^.

In summary, a global multi-omic analysis of the peripheral immune cell compartment supports that more severe cognitive impairment in AD is associated with a shift towards an activated peripheral innate immune profile accompanied by lower B cell frequencies.

### Low-grade inflammatory programming defines expanded classical monocyte compartment in Alzheimer’s Disease

As monocytes exhibited significant alterations in multiple modalities on a global scale, we further evaluated their phenotype in greater detail (Figure 2A). Based on the expression of CD14 and CD16, we annotated a total of 4.3 million cells (downsampled to 387,144) in the MCFC monocyte space (Figures 2B, S2A), revealing a significant increase of classical monocytes uniquely in ADD patients compared to controls, with a simultaneous decrease in non-classical monocytes, independent of age within groups (Figures 2C, S2B). To identify stable transcriptional changes in the scRNA-seq subset of 92,111 monocytes, we performed differential gene expression analysis on the compartment of classical (*CD14+*) monocyte states identified by subclustering, while amending for donor heterogeneity (see methods, Figures 2D-E, S2C). The highest number of differentially expressed genes (DEGs) was detected when comparing ADD to controls (Figure 2E). Signature enrichment analyses showed a gradual shift of these gene expression patterns along the AD trajectory, while they were not driven by age, sex or ApoE genotype (Figure S2D-E, Table S2). This was underlined by common gene expression patterns in ADD and MCI including genes like *FOSB*^32^, *FCAR*^33^, *ABCA1*^34^, *AREG*^35^, *KLF4*^36^, *KLF6*^37^, and *NR4A3*^38^, all associated with inflammatory processes (Figure 2F). Notably, *FOLR3* was among the commonly downregulated genes in ADD and MCI, a gene that plays a role in downregulating the AD risk factor homocysteine^39^.

**Figure 2 –.**
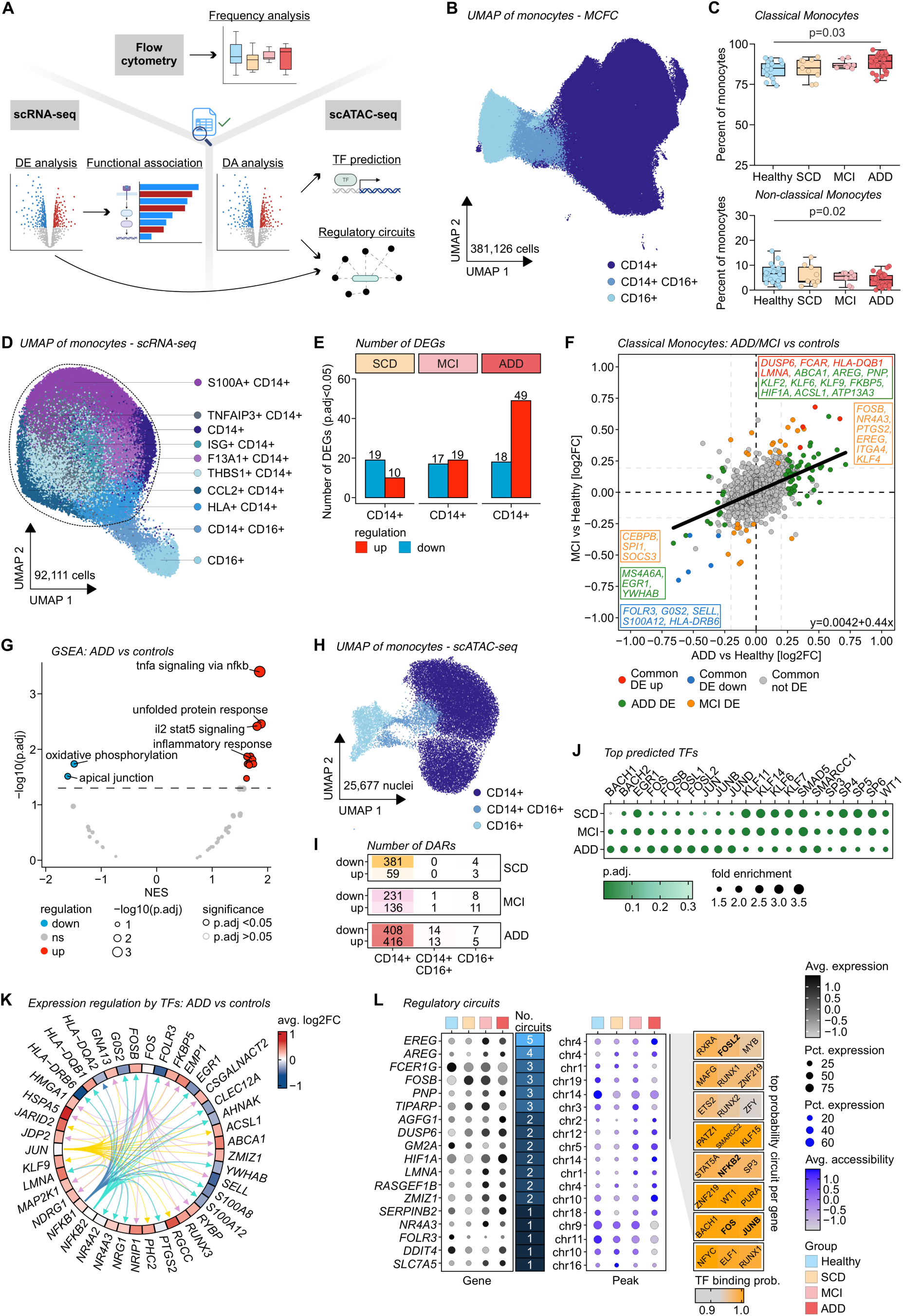
Low-grade inflammatory programming defines expanded classical monocyte compartment in Alzheimer’s Disease. (A) Schematic overview of the cell type-wise analysis framework: Assessment of (1) frequencies in MCFC, (2) differential expression (DE) analysis combined with functional association and transcription factor (TF) prediction in scRNA-seq and (3) TF prediction and regulatory circuit analysis based on differential accessibility (DA) analysis in scATAC-seq. Created with BioRender. (B) UMAP visualization of the monocyte compartment in the MCFC data colored by cell state (n = 4,294,670 cells were downsampled to 387,144 cells for visualization). (C) Relative frequencies of classical (CD14+) and non-classical (CD16+) monocytes in MCFC as percentage of total monocytes. Statistical analysis using Kruskal-Wallis and post hoc Dunn’s test, Benjamini-Hochberg-adjusted p-values are indicated. (D) UMAP visualization of the monocyte compartment in the scRNA-seq data, colored by cell state (n = 99,111 cells). Dashed line circles classical (*CD14+*) monocytes. (E) Number of differentially expressed genes (DEGs) in classical monocytes per A+ group compared to controls colored by regulation. Differential expression defined by Bonferroni-adjusted p-value < 0.05 and absolute average log2 fold change > 0.2 (iterative logistic regression framework with correction for season of sampling and sample of origin). (F) Fold change comparison of genes in classical monocytes for ADD and MCI against the control group, colored by regulation (min.pct > 0.1). (G) Gene set enrichment analysis (GSEA) based on hallmark database terms for classical monocytes comparing the ADD and control group. NES = normalized enrichment score. (H) UMAP visualization of the monocyte compartment in the scATAC-seq data colored by cell state. (I) Number of differentially accessible regions (DARs) compared to controls in monocyte states (n = 25,577 nuclei). (J) Top enriched factors from transcription factor (TF) prediction based on the scATAC-seq data per group. Dot size indicates fold enrichment of the predicted TF in the respective group; dot color intensity indicates adjusted p-value. (K) Circus plot depicting selected TFs predicted from the scRNA-seq data in classical monocytes and their potential target genes colored by fold change between the ADD and control group. (L) Regulatory circuit prediction of DEG-DAR-TF based on combined analysis of the scRNA-seq and scATAC-seq data using the tool MAGICAL. For peaks, the respective chromosome is indicated. TFs for the first 8 circuits are shown.

Among all nine monocyte states revealed by subclustering (Figures 2D, S2C), a *CCL2*+ *CD14+* classical state was overrepresented in ADD and depicted the most prominent enrichment of the ADD DEG set, indicative again of an inflammatory reprogramming of monocytes in ADD^40^ (Figure S2F-G). GSEA and functional enrichment of DEGs in classical monocytes elucidated “*TNFa signaling via NFkB”* and “*Inflammatory response”* as top upregulated terms for ADD and MCI further supporting the notion of low-grade inflammation (Figures 2G, S2H). Using activity inference (see methods), we additionally predicted the NF-*k*B pathway as highly active, particularly in both MCI and ADD compared to controls and SCD (Figure S2I). “*TNFa signaling via NFkB”*-related DEG expression patterns underscored the gradual development of the phenotype in the early AD trajectory (Figure S2H, S2J). Further investigation of the downregulated genes revealed an association with oxidative phosphorylation (Figures 2G, S3A).

To unveil potential epigenetic regulatory mechanisms encoding for the observed alterations along the AD trajectory, we subclustered and annotated a total of 25,677 nuclei in the scATAC-seq monocyte space (Figures 2H, S3B) and confirmed the increase in classical monocytes in ADD (Figure S3C). A refined differential accessibility (DA) analysis confirmed major changes in accessibility for classical monocytes particularly in ADD (408 down- and 416 upregulated differentially accessible regions (DARs)) (Figure 2I). Transcription factor (TF) prediction based on the DARs of all A+ groups pointed to members of the AP-1 complex as key regulators across all stages (Figure 2J), indicating an epigenetic contribution to the observed inflammatory reprogramming during AD. Motif enrichment for these TFs supported a specific enrichment of *FOS* in ADD and MCI (Figure S3D), as well as the enrichment for *NFKB1*, as predicted from the scRNA-seq data (Figure S2I).

To further validate this finding, we conducted TF prediction based on the presence of binding motifs (TFBS) adjacent to DEG loci in the scRNA-seq data. AP-1 and NF-*k*B-related TFs (*FOS, JUN, NFKB1* and *NFKB2*) were among the top predicted regulators for the DEG sets derived from the ADD and MCI groups (Figure 2K). Genes of the NF-*k*B and AP-1 TF complexes, like *NFKB1, NFKB2, ATF3, and ATF4* also showed increased gene expression in the ADD and/or MCI group, respectively (Figure S3E). ChromVAR analysis further supported increased accessibility not only in *FOS* and *NFKB1* gene bodies but also in *FOS* and *NFKB1* motif-rich loci along the AD trajectory (Figures S3D, S3F).

Combining the information from chromatin accessibility and gene expression data to predict potential disease-associated regulatory circuits using Multiome Accessibility Gene Integration Calling and Looping (MAGICAL, see methods)^41^ (Figure 2L), we identified 69 candidate peak-gene-TF circuits in ADD associated with 38 genes (Figure S3G). We prioritized genes associated with more than one circuit, hypothesizing that these provide more relevant contributions to the overall multi-layer regulation (Figure 2L). The analysis revealed FOS proteins as major candidate regulators, as not only increased expression of *FOSB* was found associated with 3 circuits, but FOS and FOSL2 were also predicted as top potential TFs regulating some of the most recurrent circuit-associated genes. The analysis also pointed towards a role for amphiregulin (AREG) and epiregulin (EREG), two EGF-like growth factors modulating monocyte but also neuronal functions^42,43^, which showed higher expression in ADD or MCI (Figures 2F, S2J).

As we observed genes associated with known genetic AD risk loci among the DEGs in ADD (e.g. *ABCA1*^34,44^), we assessed their expression differences in A+ groups in classical monocytes. Multiple risk genes depicted significant up- or downregulation in ADD (32.6% of risk genes), MCI (29.3%) and SCD (10.9%) (Figure S3H), indicating altered expression of these genes in classical monocytes in relation to the AD trajectory.

Taken together, we observed a reprogramming of an expanded classical monocyte compartment along the AD trajectory. This reprogramming involved both transcriptional changes evidenced by a low-grade inflammatory signature and epigenetic modifications mediated largely by AP-1 and NF-*k*B activity with prodromal MCI sharing transcriptional and epigenetic patterns with ADD.

### Dysfunctional, terminally differentiated CD8+ T cells expand in Alzheimer’s Disease

Previous studies reported an expansion of terminally differentiated effector memory re-expressing CD45RA+ (Temra) CD8+ T cells in CSF and peripheral blood of patients with amyloid-associated cognitive dysfunction and suggested a potential involvement in disease progression^45^. Building on this, we assessed the T cell compartment in more detail as we had observed alterations at the global scale associated with cognitive decline in AD (Figure 1H). Subclustering and annotation based on expression of CCR7 and CD45RA in the MCFC space of a total of 2 million CD8+ T cells (downsampled to 376,330 cells) (Figure 3A, S4A) showed a significantly higher frequency of Temra cells in ADD patients compared to controls, while numbers of naive (Tn), central memory (Tcm), effector memory (Tem) were not significantly different (Figures 3B, S4B).

**Figure 3 –.**
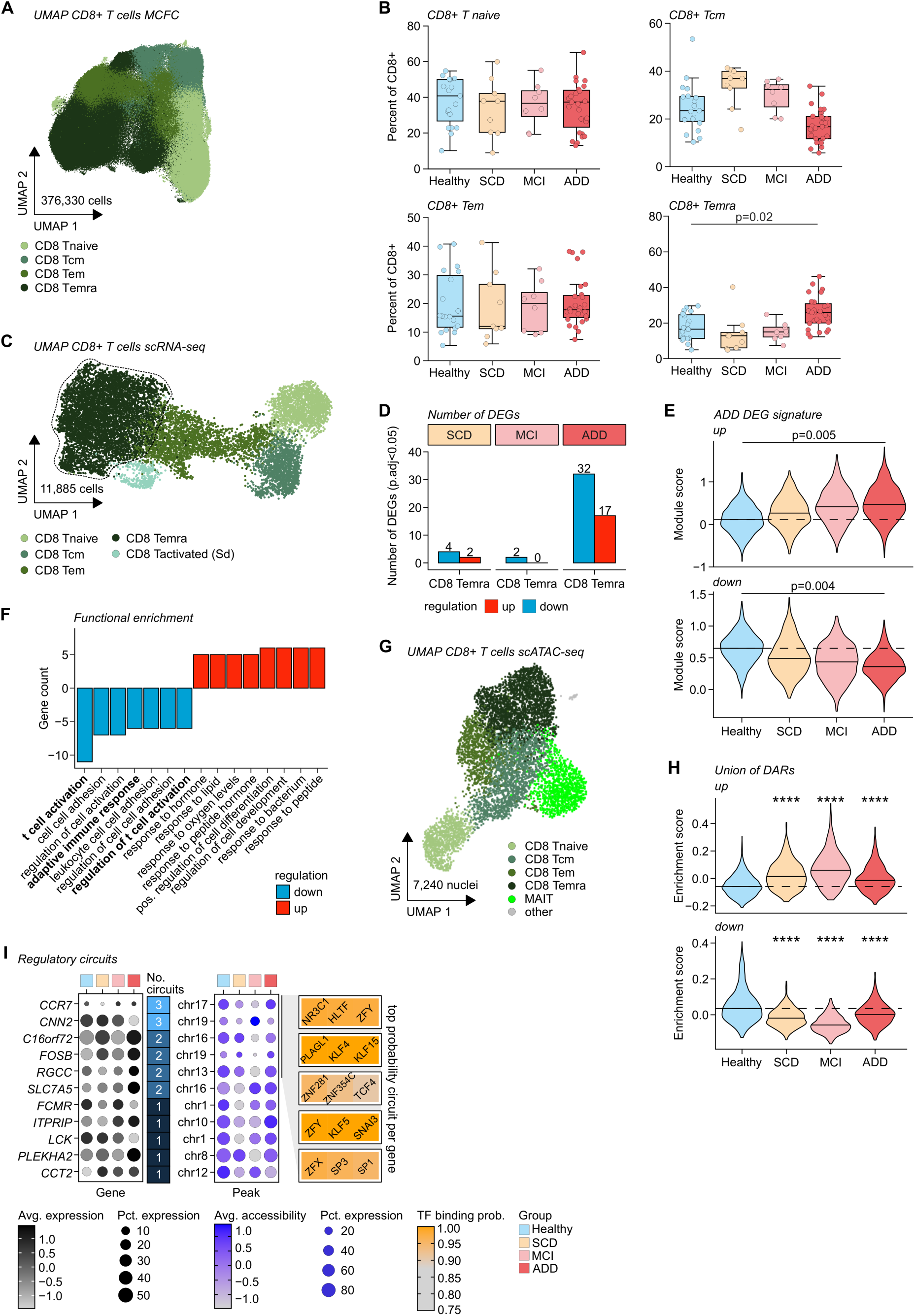
Dysfunctional, terminally differentiated CD8+ T cells expand in Alzheimer’s Disease. (A) UMAP visualization of the CD8+ T cell compartment in the MCFC space colored by cell state (n = 1,991,302 cells were downsampled to 376,330 cells for visualization). (B) Relative frequencies of CD8+ T cell states as percentage of total CD8+ T cells. Statistical analysis using Kruskal-Wallis and post hoc Dunn’s test, Benjamini-Hochberg-adjusted p-values are indicated. (C) UMAP visualization of CD8+ T cells in the scRNA-seq dataset colored by cell state (n = 11,885 cells). Dashed line circles CD8+ Temra cells. (D) Number of differentially expressed genes (DEGs) in CD8+ Temra cells per A+ group compared to controls colored by regulation. Differential expression defined by Bonferroni-adjusted p-value < 0.05 and absolute average log2 fold change > 0.2 (iterative logistic regression framework with correction for season of sampling and sample of origin). (E) Module score of ADD-derived DEG signatures per group for the CD8+ Temra cell space in the scRNA-seq dataset integrated for season of sampling. Statistical analysis using Wilcoxon test on sample level and Benjamini-Hochberg-adjusted p-values are indicated. (F) Functional enrichment of GO biological processes (BP) database terms based on DEGs in CD8+ Temra cells colored by regulation. Top terms by gene count are depicted. (G) UMAP visualization of CD8+ T cells in the scATAC-seq dataset (n = 7,240 nuclei). (H) Enrichment of the union of differentially accessible regions (DARs) in the scATAC-seq dataset. Statistical analysis using Wilcoxon test. **** indicates p-value < 0.0001. (I) Regulatory circuit prediction of DEG-DAR-transcription factor (TF) based on combined analysis of the scRNA-seq and scATAC-seq data using the tool MAGICAL. For peaks, the respective chromosome is indicated. TFs for the first 5 circuits are shown.

Gene expression patterns in CD8+ T cells were examined in a total of 53,733 T cells (Figures 3C, S4C-E) and confirmed the overrepresentation of the Temra subset in the ADD group, which also showed the highest number of DEGs (Figures 3D, S4F). This transcriptional profile reflected a state driven by later-stage AD processes with functional and gene set enrichment analyses revealing a consistent downregulation of T cell activation-related genes in ADD compared to controls, which already partially manifested in SCD and MCI, largely independent of age (Figures 3D-F, S4G-H).

To explore whether the dysregulation of CD8+ T cell function was accompanied by epigenetic changes, we assessed the scATAC-seq data, subclustering 7,240 CD8+ T cells into Tn, Tcm, Tem, and Temra cells (Figures 3G, S5A). DAR analysis identified the highest number of DARs in Temra cells from MCI patients (52 downregulated, 13 upregulated) (Figure S5B) and module enrichment of the union of up- and downregulated DARs revealed significant differences across the AD trajectory (Figure 3H), supporting a concept where the epigenetic changes precede and drive the accumulation of Temra cells and their transcriptional alterations in ADD patients. This was further underlined by rank-gene-based functional analysis pointing towards modulation of inflammation-related responses in Temra cells from MCI and SCD patients (Figure S5C). To identify potential disease-modulating peak-gene-TF circuits in the CD8+ Temra cell compartment, we combined gene expression and chromatin accessibility information, revealing key genes such as *CCR7* and *CNN2* that were associated with three circuit motifs indicative of altered adaptive immune function in ADD (Figure 3I). CCR7 is a known regulator of T cells migration into the CNS as well as a modulator of neuroinflammation associated with worsened glymphatic function, cognition, and Aβ pathology^46,47^. Calponin-2 (CNN2) regulates actin dynamics and migration, and expression quantitative trait loci (eQTLs) in the *CNN2* gene locus have been described for AD both in blood and brain tissue^48^.

Within the CD4+ T cell compartment, we observed no overrepresentation of any subset in any A+ group (Figure S5D-F). On a functional level, CD4+ T cells from ADD patients displayed upregulation of genes linked to innate immune and stress processes as in monocytes, along with downregulation of genes associated with activation as in CD8+ T cells (Figure S5G-H). At the chromatin level, functional enrichment of specific DAR-associated genes also highlighted an upregulation of stress response genes as well as an overall regulation of cell activation in the A+ groups, while no changes in cell state frequencies were observed (Figure S5A, I-K). Overall, the CD4+ T cell compartment shared some ADD-related characteristics with the CD8+ T cells, though less prominently.

In summary, we observed that the AD trajectory is associated with a perturbed T cell compartment. The most striking changes were in CD8+ T cells, particularly expansion and dysregulation of CD8+ Temra cells marked by both transcriptional downregulation of activation-related genes and epigenetic alterations linked to inflammatory responses.

### Contraction and loss of function define B cell immunity in Alzheimer’s Disease

We investigated how the B cell compartment is affected in the AD trajectory given the overall reduced frequency of B cells (Figures 1F, S1E). Subclustering the MCFC dataset, we analyzed over 2.3 million B cells and delineated four major subsets including naïve, non-class-switched, class-switched, atypical B cells and plasmablasts (Figures 4A, S6A). We observed that the frequency of class-switched memory B cells was significantly decreased in ADD patients, independent of age (Figure 4B, S6B).

**Figure 4 –.**
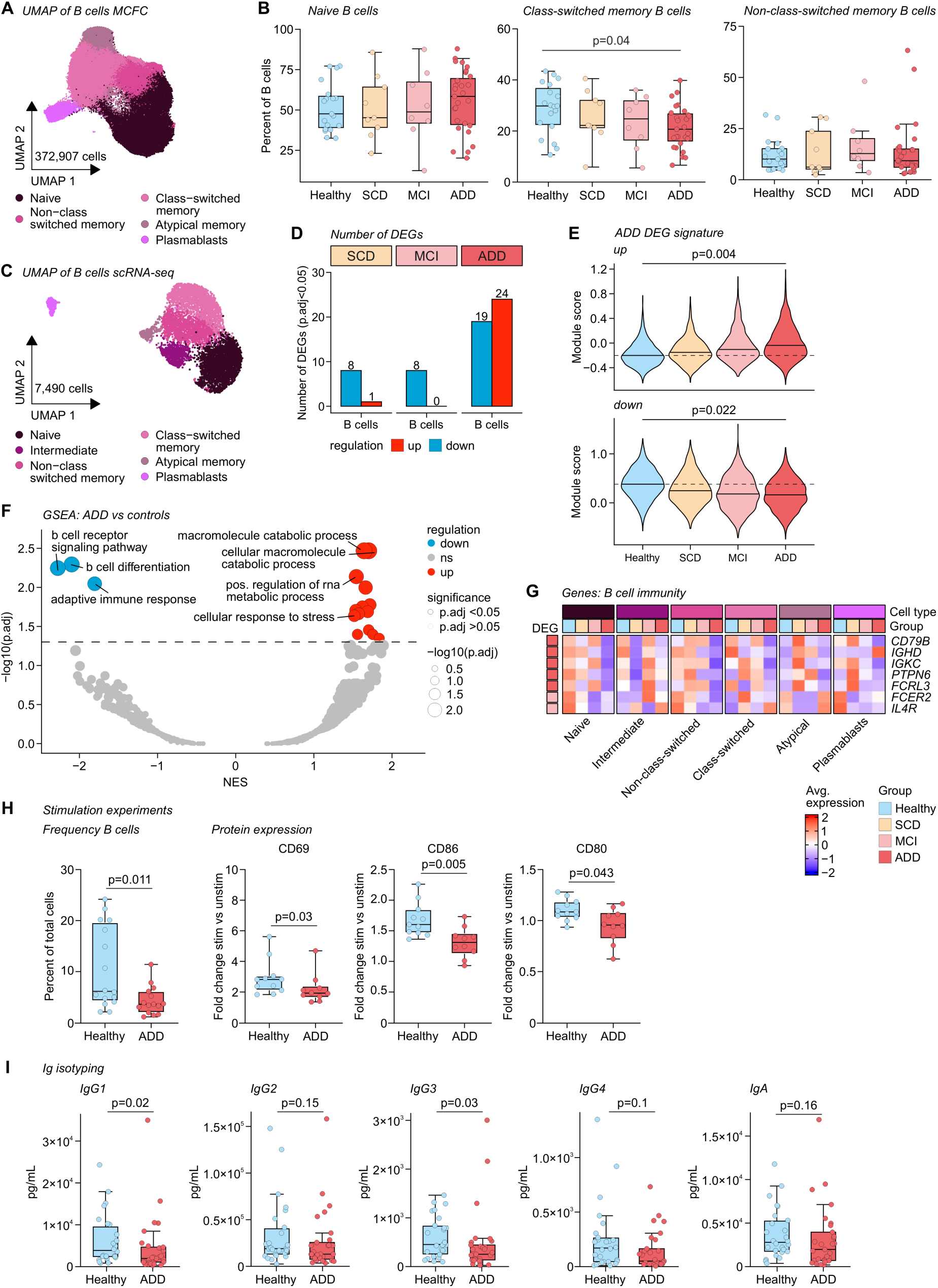
Contraction and loss of function define B cell immunity in Alzheimer’s Disease. (A) UMAP visualization of the B cell compartment in the MCFC data colored by cell state (n = 2,253,572 cells were downsampled to 372,907 cells for visualization). (B) Relative frequencies of B cell states as percentage of total B cells. Statistical analysis using Kruskal-Wallis and post hoc Dunn’s test, Benjamini-Hochberg-adjusted p-values are indicated. (C) UMAP visualization of the B cell compartment in the scRNA-seq data, colored by cell state (n = 7,490 cells). (D) Number of differentially expressed genes (DEGs) in B cells per group compared to controls colored by regulation. Differential expression defined by Bonferroni-adjusted p-value < 0.05 and absolute average log2 fold change > 0.2 (iterative logistic regression framework with correction for season of sampling and sample of origin). (E) Module score of ADD-derived DEG signatures per group for the B cell space in the scRNA-seq dataset integrated for season of sampling. Statistical analysis using Wilcoxon test on sample level and Benjamini-Hochberg-adjusted p-values are indicated. (F) Gene set enrichment analysis (GSEA) on GO biological processes (BP) in B cells comparing the ADD and control group. NES = normalized enrichment score. (G) Heatmap of average expression of DEGs related to B cell immunity in the different groups per cell state. (H) B cell-specific read-out from PBMC stimulation experiments. Cells were stimulated with LPS and measured by flow cytometry. Percentages of total CD45+ cells or stimulation-induced fold changes in mean fluorescence intensity (MFI) for CD69, CD80, CD86 are shown. Statistical analysis using Wilcoxon test or Student’s t-test, p-values are indicated. (I) Flow cytometric read-out from immunoglobulin isotyping assay in predicted pg/mL. Statistical analysis using Wilcoxon test, p-values are indicated.

To gain insights into the transcriptional profiles demarcating A+ groups from controls, we subclustered all B cells in the scRNA-seq data. Aligning with the MCFC data, the analysis revealed the four described subtypes and one cluster of intermediate cells (Figure 4C, S6C-D). DE analysis revealed that ADD exhibited the most prominent changes in gene expression, not majorly driven by demographic factors or ApoE genotype (Figure 4D, S6E-F, Table S2). In contrast, SCD and MCI showed less prominent alterations compared to controls (Figure 4D), with partial overlap in the enrichment of the ADD-derived DEG sets (Figure 4E).

GSEA highlighted a negative enrichment for pathways involved in B cell receptor signaling, differentiation, and adaptive immune responses in ADD (Figure 4F), pointing towards impaired B cell function. Additionally, similar to what we observed in the T cell and monocyte compartments, stress-response pathways were positively enriched in ADD. Key downregulated DEGs associated with B cell function, activation or receptor signaling (*CD79B*^49^*, IGHD*^50^*, IGKC*^51^*, PTPN6*^52^*, FCRL3*^53^*, FCER2*^54^, and *IL4R*^54^) exhibited a similar expression pattern across all major B cell subtypes for ADD patients (Figure 4G), highlighting an overall suppressed B cell activation state in ADD further supported by the loss of class-switched memory cells (Figure 4B, S6G).

To validate these transcriptional findings at the functional level, we assessed immediate B cell activation in samples independent of the discovery cohort by measuring the upregulation of canonical activation markers (CD80, CD86, and CD69) following *invitro* stimulation of PBMCs with lipopolysaccharide (LPS). This analysis revealed significantly diminished upregulation of these activation markers upon stimulation indicating a limited activation potential of the B cell compartment (Figure 4H). Moreover, we conducted immunoglobulin (Ig) isotyping on serum samples and found significantly reduced concentration of class-switched isotypes in ADD patients compared to controls, reinforcing the concept of a reduction of memory B cells and humoral immune potential in ADD (Figure 4I).

The comprehensive investigation of the B cell compartment revealed that ADD is associated with a decrease in B cells, particularly class-switched memory B cells, and linked to an impaired activation potential. Both transcriptomic alterations and functional assays underscore a compromised ability to mount effective humoral immune responses in AD.

## Discussion

AD remains a major challenge in global healthcare, despite the recent introduction of the first disease-modifying therapies for a subset of AD patients. Advances in diagnostic biomarker development including tau species like p-tau-217^55^ or Aβ have improved diagnostic precision, yet disease outcome prediction and progression as well as the role of additional molecular and cellular mechanisms require further attention to unravel the disease’s underlying mechanisms as a prerequisite for additional preventative and therapeutic options. Recent lines of evidence have increasingly pointed to immune dysregulation within the brain itself as an emerging component in the pathophysiology of AD, while the role of systemic immune deviation is incompletely understood^18,56^.

By making use of a large well-characterized cohort and a single-cell level multi-omics approach we identified alterations in peripheral immune cells that provide new insights into the systemic processes potentially contributing to AD pathogenesis. These observed changes are likely missed without the use of high-resolution profiling, but their confirmed presence using an integrative multi-omics approach depicts their particular relevance for the AD trajectory following this framework. Here, we outlined evidence for a low-grade inflammatory state regulated by AP-1 and NF*-k*B complexes within an expanded classical monocyte population in AD, a finding that is in line with previous suggestions^57^. Notably, the observed low-grade inflammatory monocyte state was paralleled by disruptions in mitochondrial function, previously documented in brains derived from AD patients post mortem^58^. This suggests a connection between inflammation, mitochondrial dysfunction and the observed reprogramming of peripheral monocytes during disease progression in the context of AD. ^5923,25,60,61^. In addition to monocyte deviations, we observed elevated levels of functionally dysregulated CD8+ Temra cells in ADD patients, which reinforces the concept of T cell dysfunction at both proportional and transcriptional level, preceded by epigenetic changes. Consistent with this, previous studies have demonstrated that the presence of CNS-invading CD8+ T cells, particularly Temra cells, is negatively related to cognitive function, and is associated with perturbation and clonal expansion of these cells^14,28,45,62^. Even though clonality cannot be assessed in our multi-omic approach, the association of adaptive immune potential to the A+ groups suggest that immune alterations evolve with AD progression.

Contrary to expansion of classical monocytes and CD8+ Temra cells, we detected a reduction in B cell frequencies, particularly within the class-switched memory subset, and an accompanying reduction of serum Igs in AD. Moreover, our analyses indicated a diminished activation potential of B cells,which may compromise their capacity to mount effective humoral responses. These findings contribute to ongoing debates regarding the dual roles of B cells in neurodegenerative contexts, as recent reports suggested both beneficial and detrimental roles for B cells and for the outcome of modulating their activity in AD^21,63–65^. Our findings on adaptive immune cells suggest a greater benefit for AD patients from recurrent vaccinations to decrease infection susceptibility, especially considering the potential contribution of infections and systemic inflammation to the aggravation of cognitive decline in AD^66^. Indeed, the beneficial effects of vaccination against shingles on AD incidence and prevalence would further support such hypotheses^66^.

Collectively, by delineating multi-layered immune alterations, our study provides evidence for peripheral immune dysregulation along the AD trajectory and highlights the potential for immunotherapeutic approaches as a future direction in disease management. Systemic immunity in AD is characterized by increased systemic stress of the innate immune system and an impaired potential in adaptive immunity. These findings not only refine our understanding of AD pathogenesis but also open avenues for exploring immunomodulatory strategies. In particular, targeted interventions to bolster immune responses might offer novel therapeutic benefits in managing AD and hold the potential for maintaining immune competence in elderly people at risk for dementia.

## Limitations of the study

Our study provided valuable insights into the peripheral immune dysregulation during the AD trajectory, revealing that innate and adaptive immune compartments are differentially disturbed in a cell type- and disease stage-dependent manner, which also indicated variable opportunities for intervention. Still, the study design was limited to the structure given by the DELCODE/DESCRIBE cohorts, which restricted our analysis to a cohort that did not allow us to draw conclusions on the influence of ApoE genotypes in great detail due to unbalanced distributions between groups.

This study provides a critical insight into the complex interplay between AD and the peripheral immune system and offers a new perspective on the potential for immunotherapies to target and correct the immune deviations that contribute to AD pathology, thereby opening up new avenues for the development of effective treatment or prevention strategies including vaccines. The exact cascade of cause and consequence between immune dysregulation and brain pathology as well as AD-enforced lifestyle of patients remains to be fully understood.

## STAR★Methods

### Key resources table

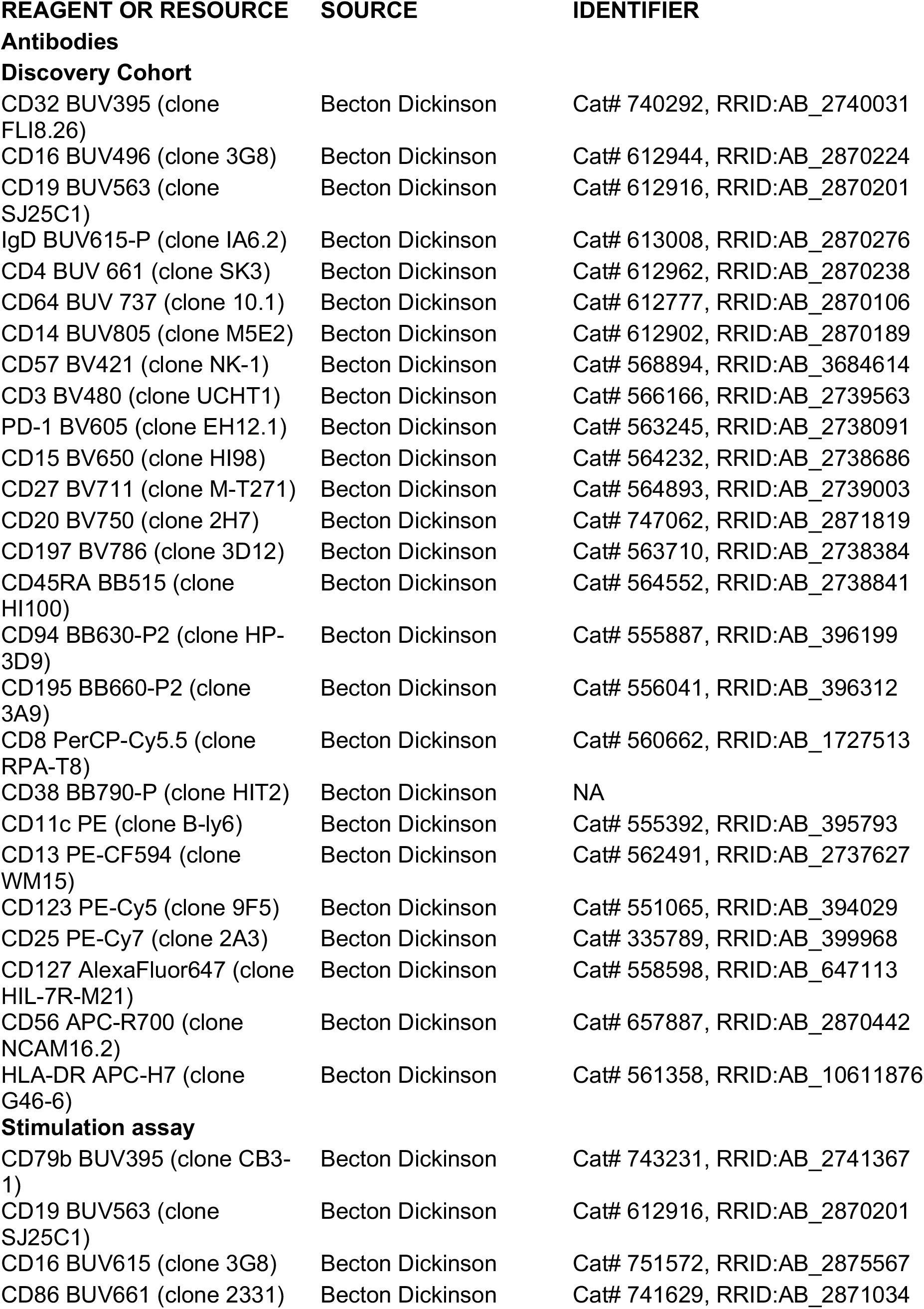
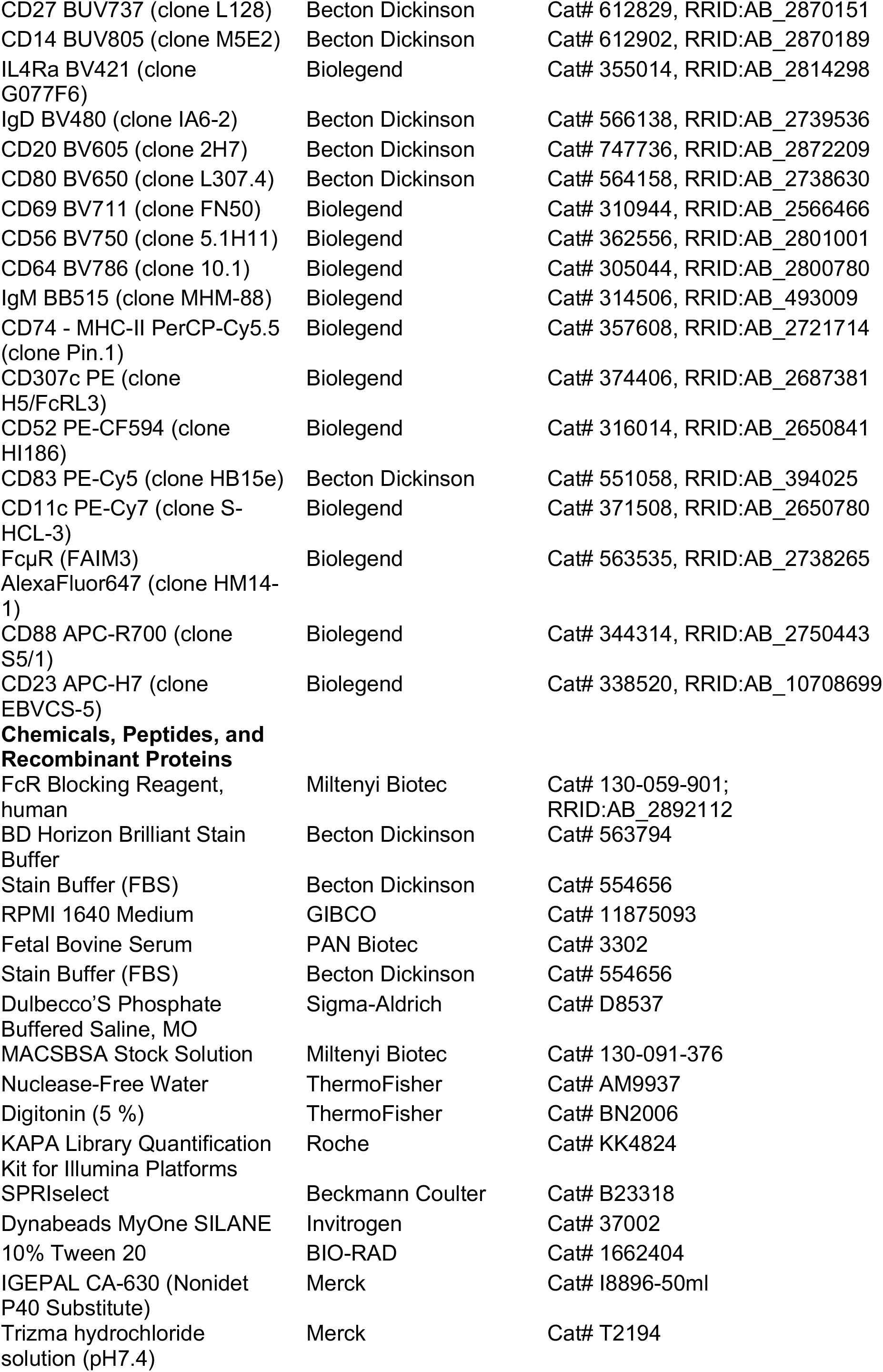
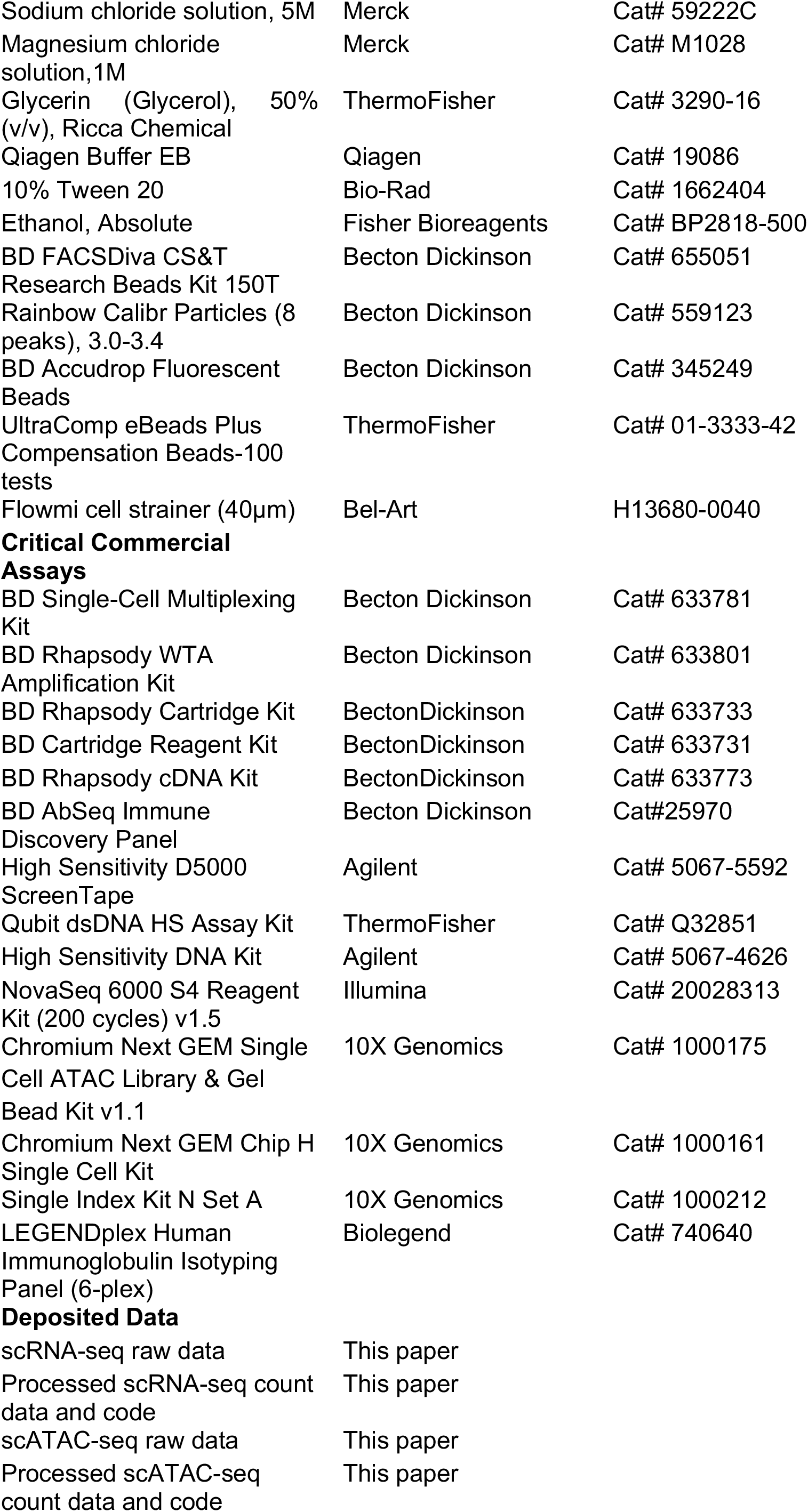
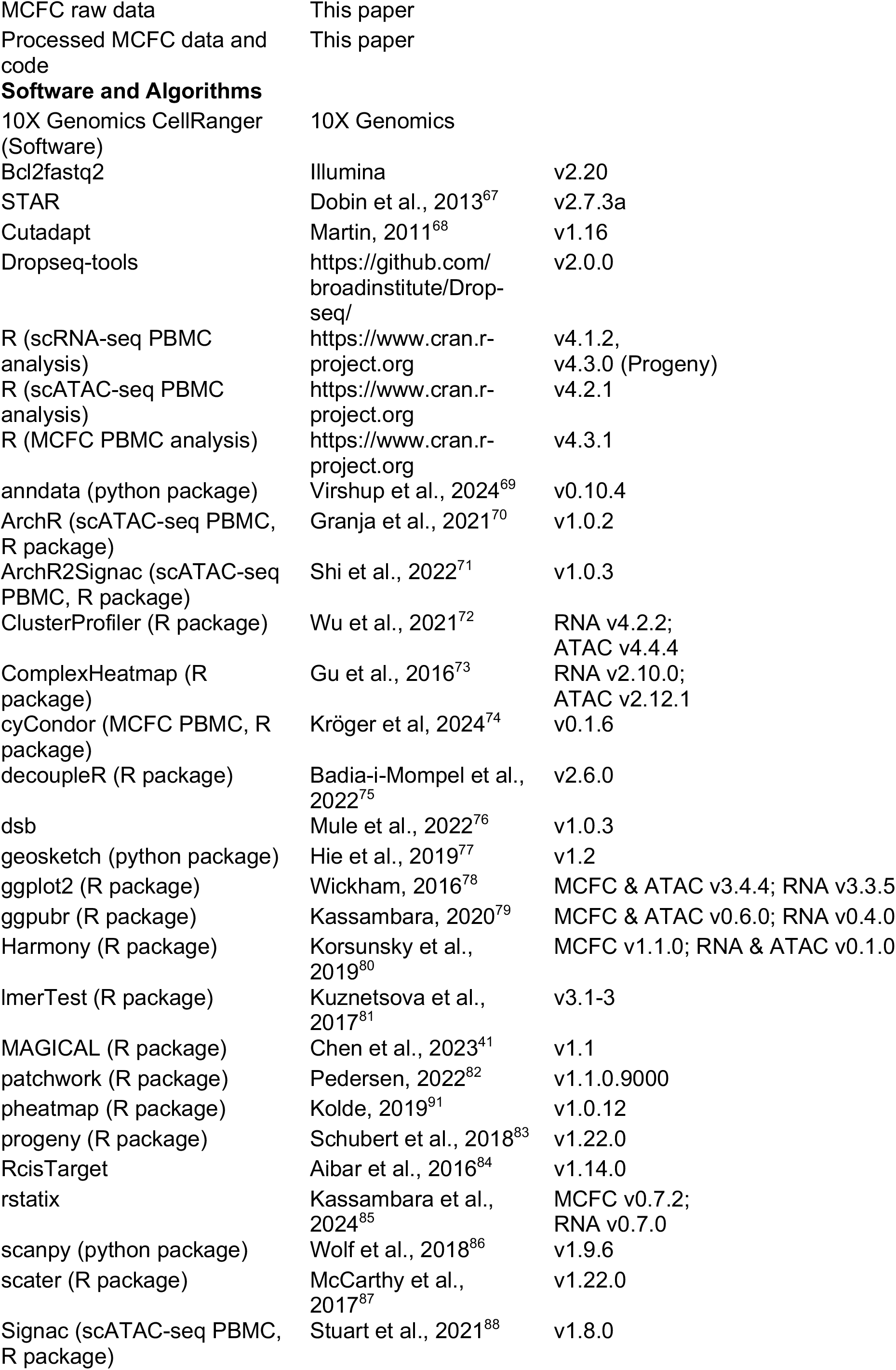
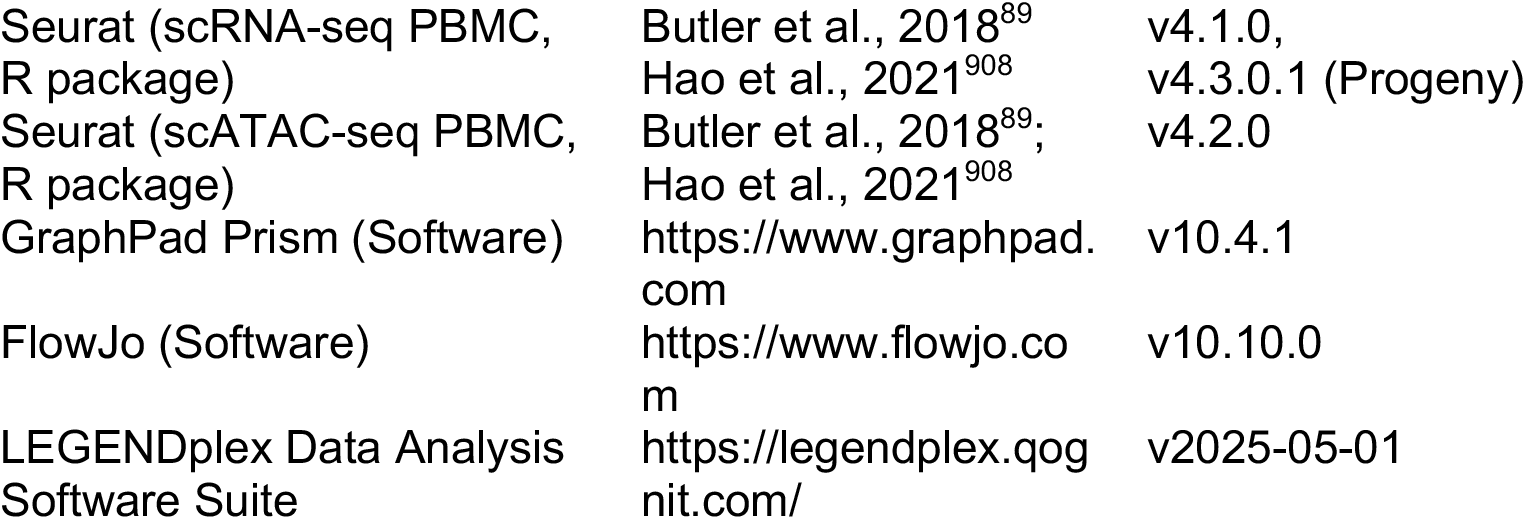

### Resource availability

#### Lead contact

Further information and requests for resources and reagents should be directed to and will be fulfilled by the lead contact, Marc D. Beyer.

#### Materials availability

This study did not generate unique new reagents.

#### Data and code availability

scRNA-seq, scATAC-seq and MCFC data are deposited at the DZNE Clinical Research Platform. Code will be deposited at Zenodo and is available as of the date of publication. Any additional information required for data reanalysis is available from the lead contact upon request.

### Experimental model and study participant details

#### Participant cohorts and study design

To identify and determine specific molecular signatures in the peripheral blood of A+ AD patients that allow the characterisation of disease stage and evaluation of disease progression, a total of 242 PBMC samples from participants older than 60 years and enrolled in the DELCODE study, an observational longitudinal memory clinic-based multicenter study conducted by the DZNE were analyzed by scRNA-seq, scATAC-seq, MCFC and proteomics. Another 26 samples were derived from participants enrolled in the DESCRIBE study of the DZNE. 170 samples of 95 participants included in the discovery cohort were selected to be roughly age- and sex-matched between groups. 75 participants were additionally sampled at a one-year follow-up time point, leading to a split of the discovery cohort in two time point subgroups. The remaining 98 available samples from 98 participants without further follow-up were used to define the internal validation cohort. All included samples were collected between November 2015 and September 2020. The final number of samples included in the subgroups of scRNA-seq, scATAC-seq, and MCFC was determined modality-wise after filtering based on CSF biomarker status (Aβ positivity) according to the ATN framework and modality-specific quality criteria, as outlined in detail in the sections below. The final selection of patients per analysis including clinical characteristics is given in Table S1.

#### Definition of clinical groups

Grouping of participants into groups of the AD trajectory and controls was based on the criteria established for the DELCODE study before^30^. All patient groups (SCD, MCI, ADD) were assessed clinically at memory centers. The Consortium to Establish a Registry for Alzheimer’s Disease (CERAD) neuropsychological test battery was used to measure cognitive function and define the participant grouping accordingly. SCD was defined by the presence of subjectively reported decline in cognitive functioning with concerns as expressed to the physician and a test performance of better than -1.5 standard deviations (SD) below the age, sex, and education-adjusted normal performance on all subtests of the CERAD neuropsychological battery. Amnestic MCI was defined by an age, sex, and education-adjusted performance below -1.5 SD on the delayed recall trial of the CERAD word-list episodic memory tests. ADD patients with mild Alzheimer’s dementia and ≥ 18 points on the Mini-Mental-State Examination (MMSE) were included in the study. The control group (Healthy) had to achieve unimpaired cognitive performance according to the same definition as the SCD group. If the patient grouping changed during the longitudinal yearly follow-up assessments of DELCODE participants, the group was updated accordingly from that time point on. Main exclusion criteria for all groups were outlined in detail for DELCODE before^30^ and included conditions interfering with participation in the study as well as certain medical conditions like major depressive episodes or psychiatric disorders, neurodegenerative disorders other than AD, vascular dementia, history of stroke or malignant disease, severe or unstable medical condition, and significant abnormalities in vitamin B12. Prohibited drugs included chronic use of psychoactive compounds, use of anti-dementia agents in SCD, MCI, and controls, and investigational drugs for treatment of dementia or cognitive impairment one month prior to entry and for the duration of the study.

Beyond neuropsychological testing, samples were stratified further based on the participants’ CSF ATN status at their baseline time point when entering DELCODE. Cutoff values used for ATN classification were Aβ42/Aβ40 < 0.09 (A+), totalTau > 470 pg/ml (N+) and pTau181 > 57 pg/ml (T+). All patient groups had to be at least A+, and all controls A- based on this definition, as denoted in Table S1. All AD biomarkers, cutoffs, as well as ApoE genotypes were determined and defined as described before for DELCODE^30^.

#### Participant details single cell transcriptomics

A total of 69 samples from 69 participants were included in the final scRNA-seq analysis of the discovery cohort. To be included samples had to pass multiple quality criteria, including ATN status as described above, transcriptome quality (number of transcripts, genes) and transcriptomic outlier status (no separate clustering behaviour on global PBMC level). If a sample from a participant in the time point 1 subgroup of the discovery cohort did not pass the quality thresholds, the respective time point 2 sample was redirected to the time point 1 subgroup. Demographic and clinical details of all participant subgroups are available in Table S1.

#### Participant details MCFC

For the discovery cohort, 66 samples from 66 participants were used in MCFC analysis. Participants were excluded if their ATN status did not match the criteria for inclusion described earlier or if quality control standards for flow cytometry were not met (<70,000 cells were acquired or fluorescence intensity was corrupted) or if outlier detection by fitting the data with robust regression and using a False Discovery Rate-based test on the residuals identified them as outliers after refitting the model. Demographic and clinical details of all participant subgroups are available in Table S1.

#### Participant details single cell ATAC

A total of 71 samples from 71 participants were included in the final scATAC-seq analysis of the discovery cohort. To be included samples had to pass multiple quality criteria, including ATN status as described above and number of nuclei per sample (> 100). If a sample from a participant in the time point 1 subgroup of the discovery cohort did not pass the quality thresholds, the respective time point 2 sample was redirected to the time point 1 subgroup. One participant was excluded in concordance with the scRNA-seq subgroup. Demographic and clinical details of all participant subgroups are available in Table S1.

#### Participant details LPS stimulation

A subset of 31 samples (Healthy n = 16, ADD n = 15) of participants from the validation cohort recruited within the DELCODE/DESCRIBE studies were used for PBMC stimulation assays with lipopolysaccharide (LPS).

#### Participant details immunoglobulin isotyping

A subset of samples from the respective first sampling time points of participants from the discovery and validation cohort recruited within the DELCODE study were used for immunoglobulin isotyping on serum and filtered to include the union of samples from the MCFC and scRNA-seq subgroups (Healthy n = 26, ADD n = 29).

#### Ethics

All participants or their representatives provided informed consent. The study protocol was approved by the ethical committees of all involved sites^30^. DELCODE and DESCRIBE were conducted in accordance with the Helsinki Declaration from 1975.

### Method details

#### Isolation of blood cells for MCFC, scRNA-seq, scATAC-seq

Trained study assistants performed the collection, processing, and storage of the PBMC samples as well as shipment to the central biorepository of the DZNE according to the SOP as described for DELCODE before^30^. Samples were cryopreserved at -150 °C and frozen PBMCs were provided by the DZNE Clinical Research Platform.

#### Thawing cryopreserved PBMC samples

The thawing procedure is based on a cell thawing protocol provided by 10X Genomics (CG00039 Rev D). In brief, cryotubes containing frozen PBMCs were thawed in a water bath at 37 °C then transferred to a 50 ml conical tube. Next, cells were gently rinsed with warm growth medium (RPMI + 10% FCS) in five sequential steps of 1:1 dilutions, then centrifuged at 300 g for 5 min. The pellet was resuspended in complete medium (RPMI + 10% FCS) and cell concentration and viability were determined. Cells were split and immediately subjected to MCFC, scRNA-seq and scATAC-seq protocols.

#### Details for single cell transcriptomics

##### BD Rhapsody scRNA-seq on PBMCs

Whole transcriptome analysis using the BD Rhapsody Single-Cell Analysis System (BD Biosciences) was performed as previously described^91^. Cells from each sample were labeled with sample tags (BD Human Single-Cell Multiplexing Kit) following the manufacturer’s protocol. Briefly, a total number of 3 x 10^5^ cells were resuspended in 500 μl of Stain Buffer (FBS) (BD PharMingen). The sample tags were added to the respective samples and incubated for 20 min at room temperature (RT). After incubation, 200 μl BD Stain Buffer was added to each sample and centrifuged for 5 min at 300 g and 4 °C. Samples were washed two more times. Subsequently, cells were resuspended in 500 μl of cold BD Stain Buffer and counted using Countess II FL Automated Cell Counter. Labeled samples were pooled equally in BD Stain Buffer. After centrifugation for 5 min at 300 g, 4°C cell pool was resuspended in 100 µl blocking buffer (95 µl BD Stain Buffer + 5 µl Fc block) and incubated at RT for 10 min. 100μl 2x AbSeq labelling master mix (BD AbSeq oligo-coupled antibodies reconstituted in 35 µl nuclease-free water + 65 µl BD Stain Buffer) were mixed with 100 µl of the pooled samples and incubated on ice for 40 min. 2 ml BD Stain Buffer was added to the pooled sample and centrifuged for 5 min at 300 g and 4 °C. Sample was washed two more times and resuspended in 300 µl Sample Buffer before counting. Labeled sample pool was prepared for cartridge loading in 1300 µl cold Sample Buffer. For each pooled sample two BD Rhapsody cartridges were loaded with approximately 50,000 cells each. Single cells were isolated using Single-Cell Capture and cDNA Synthesis with the BD Rhapsody Express Single-Cell Analysis System according to the manufacturer’s recommendations (BD Biosciences). cDNA libraries were prepared using the BD Rhapsody Whole Transcriptome Analysis Amplification Kit following the BD Rhapsody System mRNA Whole Transcriptome Analysis (WTA) and Sample Tag Library Preparation Protocol (BD Biosciences, 23-21712-00, 7/2019). The final libraries were quantified using a Qubit™ 3.0 Fluorometer with the Qubit dsDNA HS Kit (ThermoFisher) and the size-distribution was measured using the Agilent high sensitivity D5000 assay on a TapeStation 4200 system (Agilent technologies). Sequencing was performed in paired-end mode (2∗75 cycles) on a NovaSeq 6000 System (Illumina) with NovaSeq 6000 S4 Reagent Kit v1.5 (200 cycles) chemistry.

### Details for MCFC

#### Sample processing and staining for MCFC on PBMCs

PBMCs were centrifuged at 300 g for 5 min at 4°C and the supernatant was removed by plate inversion. Cells were then resuspended in 50 µl antibody master mix and incubated for 30 min at 4°C. Cells were washed twice with 200 µl MACS buffer (1x PBS with 0.5% BSA and 0.074% EDTA), centrifuging at 300 g for 5 min at 4°C between washes. The fluorophore-coupled antibodies listed in the key resources table were used to mark their respective epitopes. Stained cells were immediately acquired using either a FACSymphony™ S6 (discovery cohort, BD Biosciences) or a FACSymphony™ A5 Cell Analyzer (validation cohort, BD Biosciences). On every acquisition day, the cytometer setup was checked using 8-peak beads (BD Biosciences), and voltages were adjusted if necessary.

### Details for single cell ATAC

#### Nuclei isolation from PBMCs

Nuclei isolation from PBMC samples was performed following the manufacturers protocol by 10X Genomics (CG000169 Rev D). Briefly, a total number of 3 x 10^5^ cells were resuspended in 500 μl PBS/ 0.04% BSA after centrifugation at 300 g for 5 min at 4°C. Samples were washed one more time before resuspension in 500 μl PBS/ 0.04% BSA. The samples were passed through a 40 µm Flowmi Cell Strainer and counted using Countess II FL Automated Cell Counter. 40,000 cells were transferred to a 0.2 ml tube and centrifuged at 300 g for 5 min at 4 °C. Everything except for 5 µl was then removed from the supernatant to avoid dislodging the cell pellet. 45 µl chilled Lysis Buffer was added and samples were incubated on ice for 4 min before adding 50 µl chilled Wash Buffer and centrifuging at 500 g for 5 min at 4 °C. Samples were washed again with 45 µl chilled Diluted Nuclei Buffer and resuspended in 7 µl chilled Diluted Nuclei Buffer. Cells were counted using Countess II FL Automated Cell Counter with Propidium iodide staining (2µl nuclei suspension + 16µl Diluted Nuclei Buffer + 2µl PI (20µg/ml)).

#### 10X Chromium scATAC-seq on PBMCs

To determine the chromatin landscape of single PBMCs the Chromium Single Cell ATAC Solution System (10X Genomics) was used. scATAC-seq libraries were prepared according to the Chromium Single Cell ATAC Reagent Kits v1.1 user guide (CG000209 Ref F). Briefly, 7,000 nuclei per sample were mixed with the Transposition Mix (7µl 10X ATAC Buffer B and 10X ATAC Enzyme (3 µl)) and then incubated for 60 min at 37 °C. Nuclei partitioning into GEMs was done by using a Chromium Controller with Chip H and GEMs were incubated thereafter in a thermal cycler for 30 min with conditions following the manufacturers protocol. GEMs were broken by adding 125 µl 10X Recovery Agent at room temperature. The DNA was purified by sequential bead-based clean-up steps. First, 200 µl of 10X Dynabeads Cleanup Mix was added to the samples, incubated for 10 min and eluted on a magnetic separator retrieving 40 µl per sample. The second cleanup was performed by adding 48 µl 10X SPRIselect reagent to the samples, incubating for 5 min and elution on a magnetic separator in 40.5 µl Elution Solution. Libraries were prepared by PCR using a Single Index kit N set A (10X Genomics) and incubated under the thermal cycling conditions recommended by the manufacturer. Sequencing libraries were subjected to clean-up with SPRIselect reagent.

Libraries were sequenced at a molarity ratio proportional to the number of expected cells in each sample. Sequencing was performed on a NovaSeq6000 instrument with an S4 XP v1.5 (200 cycles) chemistry. Sequencing protocol was defined according to 10X recommendation with both Read1 and Read2 of 50 bp and two indexes of 8 and 16 bp respectively.

#### Details for immunoglobulin isotyping

Immunoglobulin isotyping was performed on serum samples stored at -80 °C using the Biolegend LEGENDplex™ Multi-Analyte Flow Assay Kit with the Human Immunoglobulin Isotyping Panel (6-plex). The assay was carried out at room temperature. 10 µl of thawed serum samples (on ice) were diluted in assay buffer in a ratio of 1:2. Based on the manufacturer’s protocol 7.5 µl assay buffer, standard or sample and beads each were added to every well in a 96-well V-bottom plate. After 2 hours shaking (800 rpm) in the dark, 180 µl wash buffer was added and the supernatant was discarded after centrifugation at 300 g for 5 minutes. Shaking for 1 h was repeated after the addition of 7.5 µl detection antibodies and 7.5 µl SA-PE was added thereafter, before shaking for 30 minutes. Samples were resuspended in 100 µl wash buffer after two additional washing steps (180 µl, 200 µl). Samples were measured on a FACSymphony™ A5 Cell Analyzer (BD Biosciences) with a flow rate of 1.0-1.5 µl/s, acquiring at least 2,000 beads per well.

#### Details for LPS blood stimulation

For the stimulation of blood with LPS, 1 x 10^5^ cells/ ml PBMCs were seeded into flat-bottom suspension plates and stimulated with a final concentration of 100 ng/ml LPS or complete medium (RPMI + 10% FBS) and incubated at 37 °C for 24h. Plates were centrifuged at 300 g for 5 min at RT and supernatant was collected. Cells were washed with 200 µl warm PBS containing 5 mM EDTA and incubated for 20 min at 37 °C. After resuspension, cells were transferred to FACS tubes for staining, which was performed as described before.

### Quantification and statistical analysis

#### scRNA-seq analysis

##### Data pre-processing of BD Rhapsody scRNA-seq data

After demultiplexing of bcl files using Bcl2fastq2 v2.20 from Illumina and quality control, paired-end scRNA-seq reads were filtered for valid cell barcodes using the barcode whitelist provided by BD. Cutadapt v1.16 was then used to trim NexteraPE-PE adaptor sequences where needed and to filter reads for a PHRED score of 20 or above^68^. Then, STAR 2.7.3a was used for alignment against the Gencode v33 reference genome^67^. Dropseq-tools v2.0.0 were used to quantify gene expression and collapse to UMI count data (https://github.com/broadinstitute/Drop-seq/). For hashtag-oligo based demultiplexing of single-cell transcriptomes and subsequent assignment of cell barcodes to their sample of origin the respective multiplexing tag sequences were added to the reference genome and quantified as well. Specific AbSeq oligo sequences were also added to the reference genome and quantified as well.

##### PBMC scRNA-seq & AbSeq quality control

The analysis of the scRNA-seq data was performed using the Seurat v4.1.0 pipeline^89,90^ in R4.1.2. Inflection points were calculated for the distribution of unique molecular identifiers (UMIs) and genes per cartridge, and the data was refiltered for cells with these thresholds. Only genes expressed in at least 5 cells were considered further. Demultiplexing was performed in a cartridge-wise manner using the Seurat HTODemux function with a positive quantile of 0.99 and only singlets were retained. After merging the data from all cartridges singlets with more than either 30% (discovery cohort) or 35% (validation cohort) of mitochondrial (MT-) gene counts for the discovery and validation cohort, respectively, or more than 1% of hemoglobin (HBB/HBA) gene counts were excluded together with small contaminating clusters or doublets later during downstream analysis. Moreover, cartridges with substantially low number of UMIs and genes per cell were excluded from the analysis. After quality control and filtering PBMC transcriptomes were used in downstream analysis.

AbSeq protein expression data was normalized and background noise-corrected using the R package dsb v1.0.3 in a cartridge-wise manner to account for non-specific antibody binding^76^. As four cartridges for the validation dataset depicted substantially low Abseq counts, normalized AbSeq values were imputed from the remaining cells using Seurat reference mapping to ensure equally weighted AbSeq data across cartridges in downstream multimodal clustering and to avoid subsequent corrections.

##### Downstream processing and cell annotation

After quality control, the dataset was normalized, scaled, and dimensional reduction was performed using Seurat on the RNA assay. Gene counts of every cell were divided by the total counts of the cell and multiplied by a scale factor of 10,000, followed by natural-log transformation. The normalized data was used for the identification of the 2000 most variable genes using variance-stabilizing-transformation (vst). The expression data of the most variable genes were scaled, centered and used for principal component analysis (PCA). To adjust for donor variability the harmony v0.1.0 algorithm was applied^80^.

For total PBMCs, T cells, and B cells, endorsing the multimodality per cell, neighbors were calculated using Seurat’s default function FindMultimodalNeighbors with the harmony-corrected principal components (PCs) and the normalized AbSeq values, accounting for the reduced dimensionality of the BD Immune Discovery Panel (IDP). For two-dimensional representation, uniform manifold approximation and projection (UMAP) was calculated based on the multimodal nearest neighbors. Cells were afterwards clustered by the Smart-Local-Moving algorithm on the multimodal SNN graph^92^. For cell subsets (monocytes) with no advantage of multimodal clustering, neighbors were only calculated based on the harmony-corrected PCs and clustering was performed by the Louvain algorithm based on the SNN graph. The numbers of selected principal components based on an elbow plot visualization and resolutions for clustering per cell subset are depicted in Table S5.

Cluster marker genes were calculated with the Wilcoxon rank-sum-test using Seurat’s FindAllMarkers function. Genes with a log-fold change greater than 0.25, expressed in at least 20% of cells in one tested group and 10% difference in the fraction of detection between groups, were considered together with literature-derived canonical PBMC cell type marker genes and normalized IDP protein expression values per cluster to annotate and eventually summarize clusters. Samples depicting clear outlier status and aberrant gene expression patterns upon dimensionality reduction and clustering at the PBMC level were removed from the respective dataset and the procedure above was repeated on the cleaned dataset. Full cluster marker gene lists and AbSeq IDP markers, as well as their expression are provided in Table S6.

##### Differential expression analysis

Differential expression (DE) tests between the groups of interest and controls in the respective cell subsets of the scRNA-seq data were performed by the Seurat FindMarkers function using a logistic regression (LR) framework incorporating “main season of sampling” as latent variable. Genes with an absolute log-fold change greater than 0.2, at least 10% expressed in tested groups (min.pct >0.1) and with a Bonferroni-corrected p-value <0.05 were considered significantly differentially expressed genes (DEGs).

To account for sample-driven DEGs, an iterative DE approach was used that performed n iterations using n-1 samples per iteration with n being the total number of samples in the respective comparison. Only genes that occurred in n iterations were considered as DEGs. Individual-wise DE comparisons between sampling time points used a Wilcoxon rank-sum-test-based framework without latent variable. Full DEG lists per cell subset are provided in Table S7.

##### Functional and gene set enrichment analysis

Overrepresentation analysis on the DEGs was performed on the gene sets from the Gene Ontology (GO) biological process (BP)^93^ and Kyoto encyclopedia of genes and genomes (KEGG)^94^ databases, as well as the Hallmark gene sets^95^ using the R package clusterProfiler (v4.2.2)^72^. If not indicated otherwise, terms with an adjusted p-value < 0.1 (Benjamini-Hochberg adjustment) were considered further. For the term *“TNFa signaling via NFkB”* in monocytes an unadjusted p-value < 0.05 was used to depict the enrichments of all DEG sets irrespective of DEG set size. The same cutoff was used for terms in the analysis of cognitive score-associated gene sets over time due to gene set sizes.

Gene set enrichment analysis was performed based on the aforementioned gene sets and a ranked gene list as input using the R package clusterProfiler (v4.2.2). The ranked gene lists for the groups of interest were generated by performing a DE analysis (min.pct=0.1, logfc.threshold=0, test.use=’LR’, latent.vars=’main_season’) between the group of interest and the control group and ranking resulting genes by log2 fold change. If not indicated otherwise, terms with a p-value < 0.05 (Benjamini-Hochberg adjustment) were considered significant.

##### Module score calculation and comparison

The Seurat AddModuleScore function was used with the respective gene sets mainly derived from DE tests between A+ groups and controls to calculate a per-cell enrichment score. Sex-specific genes, non-ubiquituous HLA genes, B and T cell receptor variable chain genes, and hemoglobin genes were excluded from module score calculations. If indicated, the separate scores for the up- and downregulated gene sets were eventually summarized per cell for the generation of a combined module score: total score = score up - score down. To test for significance between groups, samples with too low cell numbers have been excluded (for discovery and validation cohort > 100 for monocytes, > 30 or > 50 for T and B cells), per-cell scores have been aggregated on sample level by taking the median and Wilcoxon-rank-sum-tests between groups have been performed on sample-level to avoid inflation of p-values.

##### Correlation analysis

Correlation analyses for gene set module scores with metadata variables like age were performed by first aggregating the per-cell module score as median score per sample. Subsequently the function ggscatter from the R package ggpubr v0.4.0^79^ was used to run Pearson correlation on the module scores per sample against the respective metadata variable of interest. The stat_cor function provided correlation coefficients and p-values per group of interest. For the ADAS-Cog-13 R-squared and p-values were derived from a linear model using the lm function with age and sex as covariates.

##### Pathway activity inference

NF-*k*B pathway activity was inferred using the PROGENy^83^ v1.22.0 and decoupler^75^ v2.6.0 R packages in R4.3.0. Based on the PROGENy curated collection of pathways and target genes (n = 500 per pathway) a multivariate linear model was fit for each cell using the normalized gene expression data matrix as input. The model predicted gene expression based on pathway-gene interaction weights, obtaining t-values that were interpreted as pathway activity scores.

##### Transcription factor prediction

The R package RCisTarget v1.14.0 was used for enrichment of transcription factor binding motifs in gene sets of interest^84^. The motif search was restrained to genomic regions of 10kb upstream and downstream of the respective transcriptional start sites (TSS). Motifs were annotated with the respective transcription factors and ranked by normalized enrichment score (NES) per input gene set.

##### Data integration by reciprocal PCA

To account for seasonal effects in module score calculations, cell subsets were split by main season (summer, winter) and processed separately until highly variable gene selection and PCA calculation. Integration was then performed by reciprocal PCA with Seurat’s FindAnchors and IntegrateData functions using the first 20 or 15 dimensions for monocytes or T and B cells, respectively, based on one season as a reference^96^. The per-cell module score calculation for signatures was then done on the integrated assay. For the correlation analysis between module scores and ADAS-Cog-13 in the combined dataset of validation and discovery cohorts, integration was performed the same way after merging the datasets.

##### Quantification of cell states

To compare proportions of cell states between groups of interest, cells per group were normalized to 1,000.

### Flow cytometry analysis

#### Manual gating

The FCS files were pre-processed by gating on live lymphocytes and monocytes and excluding doublets in the FlowJo software (BD, v10.10.0). Broad cell types were annotated based on their expression of canonical cell type markers. The manual gating information of broad cell types was extracted from the FlowJo workspace.

#### High-dimensional analysis and quantification of cell states

For both the discovery and the validation cohort, the fluorescence intensity values for all cells in the Singlets/Live Cells gate were exported from FlowJo (v10.10.0) and imported into R (v4.3.1) using the cyCONDOR package^74^ (v0.1.6). In R, the intensity values were then transformed using an auto-logicle transformation, with optimal width calculated for each fluorescence parameter. To obtain frequencies for each cell type as a proportion of all cells, we calculated frequencies based on a subset of 400,000 cells. To annotate more granular cell subtypes, the dataset was subset to a single main cell type. Each object was then downsampled representatively with the geosketch package^77^ For each cell type object, the autologicle-transformed values were used for PCA. To account for batch effects due to different experimental dates, the harmony batch correction algorithm implemented in cyCONDOR was applied to normalize the PCA. The normalized values of all 27 markers were then used for UMAP dimensionality reduction and Phenograph clustering. Based on expression of canonical surface markers, we annotated cell subtypes for each respective broad cell type.

Differences in cell type frequencies between groups were calculated based on the percentages of each cell type per sample and statistical differences were determined using a Kruskal-Wallis and post hoc Dunn’s test with Benjamini-Hochberg adjustment.

#### LPS blood stimulation assay analysis

The FCS files were preprocessed by gating on live B cells and excluding doublets in the FlowJo software (BD Biosciences, v10.10.0). Subsequently, B cell subsets were defined, and mean fluorescence intensity (MFI) of selected surface markers was assessed. For each sample, the fold change in MFI between the unstimulated and stimulated condition was calculated to assess differences resulting from LPS stimulation. Outlier detection was performed in GraphPad Prism (v10.4.1) and statistical comparisons between A+ groups and controls were performed using a Wilcoxon-rank-sum test or Student’s t-test.

#### Immunoglobulin isotyping analysis

Immunoglobulin (Ig) isotyping data generated with a FACSymphony™ A5 Cell Analyzer (BD Biosciences) was preprocessed using the cloud-based LEGENDplex™ Data Analysis Software Suite “Qognit” provided by the manufacturer. Resulting predicted concentrations of Ig isotypes per sample were compared between A+ groups and controls each using a Wilcoxon-rank-sum test.

### scATAC-seq analysis

#### Data pre-processing of 10X scATAC-seq data

Raw sequencing results were first converted to FASTQ files using the cellranger-atac mkfastq (v. 2.0.0) function using default setting. Result from each sample were then aligned and preprocessed with the cellranger-atac count function (v. 2.0.0) with default setting using as reference the 10x provided reference “refdata-cellranger-arc-GRCh38-2020-A-2.0.0”. Individual aligned samples were aggregated in R (v 4.2.1) using the ArchR package (v. 1.0.2) using default setting.

#### PBMC scATAC-seq quality control and downstream processing

Level 1, preprocessing and QC: Data was further processed using the ArchR framework v1.0.2^70^. Doublets were excluded using the *filterDoublets* ArchR function and high-quality nuclei were selected based on minimum TSS enrichment score of 10 and minimum 5,000 unique fragments.. Iterative LSI was performed (function *addIterativeLSI, useMatrix = "TileMatrix"* using default parameters) and dimensions highly correlated with sequencing depth were excluded. Harmony (v0.1.0) integration^80^ was applied to minimize donor-specific batch effects before clustering (resolution=0.8) and UMAP calculation. Upon manual removal of spurious clusters (donor-driven or including hetero doublets), datasets included a total of 76,061 nuclei. We then proceeded with a first-level coarse cell type annotation by both manually inspecting literature-derived cell type consensus markers and using a supervised ArchR-embedded label transfer approach (*unconstrained* followed by *constrained* integration with our scRNA-seq dataset). The same approach was applied on monocytes, T cells and CD8 compartments after subsetting and reclustering (monocytes resolution=0.4; T cells resolution=0.8; CD8+ T cells resolution=0.4) to identify more granular cell subtypes. Peaks were called grouping by main cell type using the ArchR embedded *addReproduciblePeakSet* function with default parameters and the *macs3* method, generating 312,926 peaks. For downstream evaluations, the ArchRtoSignac package v1.0.3^71^ was employed with default parameters to generate an object editable within the Signac v1.8.0^88^ and Seurat v4.2.0^89,90^ frameworks for single-cell analyses, exporting both raw peak count and gene-score matrices.

Level 2, main biological insights: Subsequent data analysis was performed within the Signac and Seurat frameworks. Data were first transformed using the *RunTFIDF* function to generate a normalized peak count matrix layer in all Signac objects. To compare proportions of cell states between groups of interest, cells per group were normalized to 1,000. DA analyses were performed using *FindMarkers* (assay = ’peaks’, test.use = ’LR’, latent.vars = ’nCount_peaks’, p.adj < 0.05, logFC = 0.2) contrasting nuclei from A+ groups (*SCD*, *MCI*, *ADD*) against *Healthy* donors within cell types and states of interest. TF enrichment on monocytes DARs was performed using the *chromVARmotifs* annotation (*human_pwms_v1*), with the *AddMotifs* and *FindMotifs* Signac functions, while *RunChromVAR* was employed to calculate deviation scores for key TF-associated motifs per nuclei across groups. The *GSEA* function (clusterProfiler v4.4.4) was employed for a log2FC gene-score matrix rank-based functional enrichment in Temra cells and key significant (p.adj < 0.05, Benjamini-Hochberg adjusted) terms were reported.The peak-gene-TF disease circuit prediction was performed using the tool MAGICAL^41^. For classical monocytes, input candidate peaks and genes included the union of DARs and DEGs called within the classical monocytes compartment across all the three disease groups (*ADD*, *MCI*, *SCD*) versus *Healthy*. The actual prediction was then performed only considering the *ADD* and *Healthy* classical monocytes cells subsets (TF-peak binding probability threshold=0.8, peak-gene looping probability threshold=0.75) and identified n=69 circuits. For Temra cells, input hits included the union of DARs and DEGs called within the whole CD8+ T cells compartment across all the three disease groups (*ADD*, *MCI*, *SCD*) versus *Healthy*. The actual prediction was then performed only considering the *ADD* and *Healthy* Temra cells subsets (TF-peak binding probability threshold=0.8, peak-gene looping probability threshold=0.75) and identified n=66 circuits.

## Statistical analysis

Statistical analyses were performed using the R package rstatix v0.7.0/v0.7.2^85^ and GraphPad Prism v10.4.1. Statistical methods are described in the figure legends or methods as appropriate.

## Data visualization

Data visualization was done using the R packages Seurat v4.1.0/ v4.2.0, cyCondor v0.1.6, ggplot2 v3.3.5/ v3.4.4, pheatmap v1.0.12, ComplexHeatmap v2.10.0, and circlize v0.4.14, as well as GraphPad Prism v10.4.1.

## Data Availability

scRNA-seq, scATAC-seq and MCFC data are deposited at the DZNE Clinical Research Platform. Code will be deposited at Zenodo and will be available as of the date of publication. Any additional information required for data reanalysis is available from the lead contact upon request.

## Acknowledgments

We thank all participants for taking part in the study and donating their blood. This work was supported by the National Dementia Strategy for Germany. MDB is supported by the Helmholtz Association and the German Research Foundation (DFG) (SFB1454 project number 432325352, IGK2168/2 project number 272482170). LB, ACA, MB, TU, JLS and MDB are members of the excellence cluster ImmunoSensation2 (EXC2151 project number 390873048).

## Author’s contributions

Conceptualization: T.U., J.L.S., M.D.B.; Investigation: J.B.S., K.M., M.v.U., J.E., E.H., J.S.-S., M.H.S., L.L., T.E., D.Ha., S.M., Y.L., I.G., C.K., J.L., T.Z., J.V.M., C.H., B.H., R.K., D.Hu., R.S., T.P., V.I., N.R., S.P., A.D., M.K., H.T., M.R., U.S.; Data analysis: J.B.S., K.M., C.C., F.E., L.H., A.F., J.E., L.B., M.D.B.; Data curation: J.B.S., K.M., C.C., M.v.U., F.E., K.Mr., E.H., S.P, M.K., H.T, L.B., M.B., M.D.B.; Resources: E.H., H.T., L.P., D.Gr., E.J.S., M.G., K.F., C.B., N.H., A.Ro., W.G., E.I.I., D.J., S.St., B.S.R., I.K., D.Go., S.So., M.Ko., S.R., M.St., I.F., A.G., M.Sc., K.B., J.H.-R., C.L., A.Ra., F.B., J.W., R.P., E.D., S.T., A.Sp., G.C.P., O.P., J.P., S.V.S., M.W., L.K., F.J., A.Sc., E.D.D, J.L.S., M.D.B; Visualization: J.B.S., K.M., C.C., T.U., M.D.B.; Supervision: M.Bu., L.B., A.C.A., M.B., T.U., J.L.S., M.D.B.; Writing - original draft: J.B.S., K.M., C.C., M.v.U., J.L.S., M.D.B.

## Supplemental information

Table S1 – Overview of included cohorts related to Figures 1-6 and S1-S6

Table S2 – Source data of enrichment scores related to Figure 2-4

Table S3 – Overview of modality- and cell subset-wise clustering resolutions related to Figure 1-4 and S1-6

Table S4 – scRNA-seq cluster marker genes and AbSeq marker expression related to Figures 1-4 and S1-6

Table S5 – scRNA-seq differentially expressed genes per cell subset (iterative LR framework and standard Wilcoxon framework) related to Figures 2-4 and S2-6

## Declaration of generative AI and AI-assisted technologies in the manuscript preparation process

During the preparation of this work the authors used large language models to optimize style of writing. Afterwards the authors reviewed and edited the content as needed and take full responsibility for the content of the published article.

---

**Figure S1 –.**
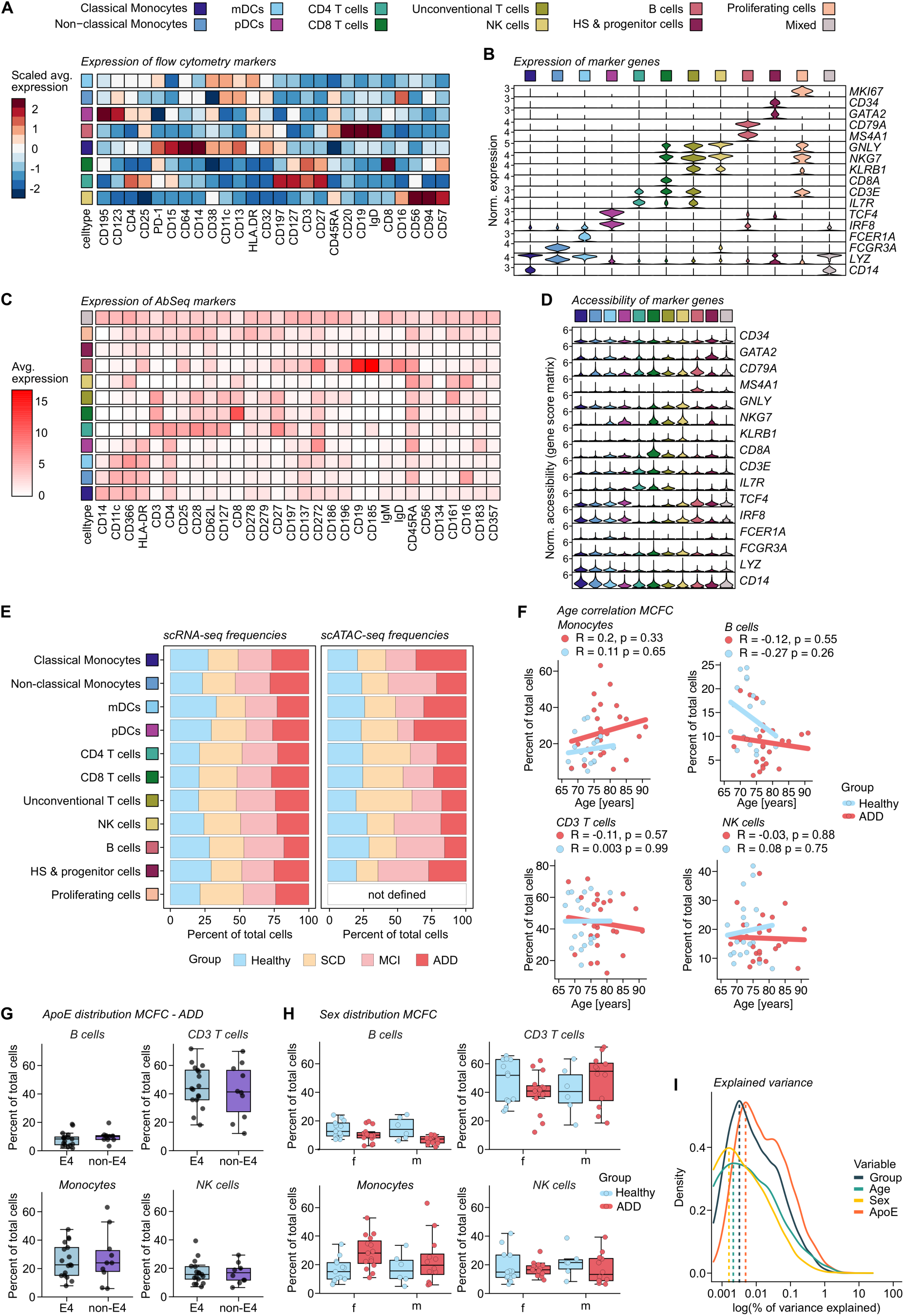
Multi-omics allow the characterization of the immune system at different stages of cognitive decline along the AD trajectory. (A) Scaled surface marker expression of all surface markers across main cell types in the MCFC dataset. (B) Normalized marker gene expression in main cell types of the scRNA-seq dataset. (C) Average dsb-normalized marker expression of the AbSeq marker panel in main cell types. (D) Normalized accessibility of marker genes of main cell types in the scATAC-seq dataset. (E) Relative frequencies of main cell types for scRNA-seq and scATAC-seq datasets across groups. Each group was normalized to 1,000 cells. (F) Linear regression between main cell type frequencies determined by MCFC and age for the control and ADD group. R (Pearson’s correlation coefficient) and p-values are indicated. (G) Relative frequencies of main cell types split by ApoE genotype within the ADD group. E4 indicates presence of ApoE ε4 allele, non-E4 indicates ε4 absence. Percentages are relative to all CD45+ live cells. (H) Relative frequencies of main cell types split by female (f) and male (m) participants. Percentages are relative to all CD45+ live cells. (I) Percentage of variance in expression of genes explained by selected variables in the sample-level metadata (group, age, sex, ApoE status).

**Figure S2 –.**
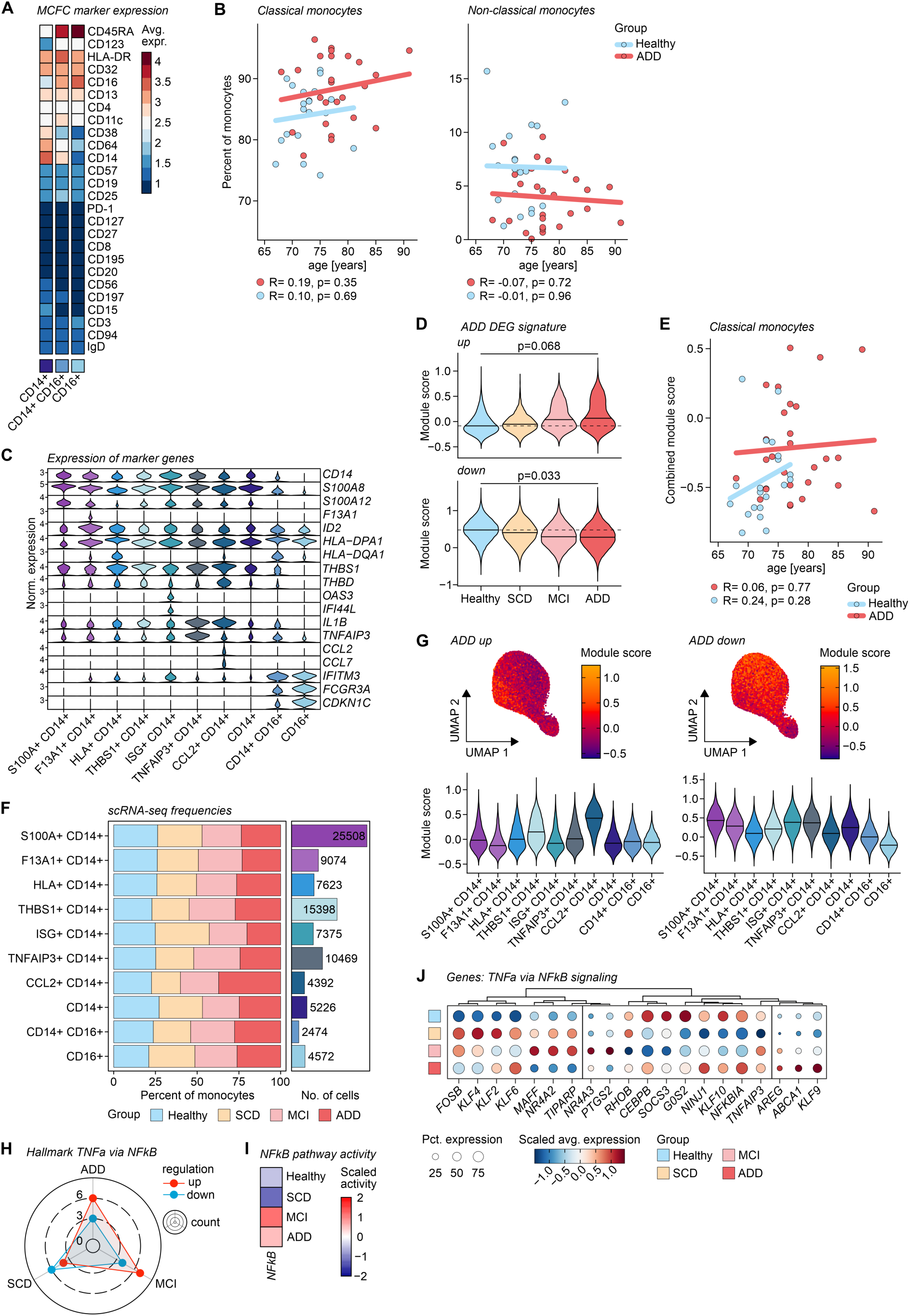
Age-independent programming of the expanded classical monocyte compartment changes along the AD trajectory. (A) Average surface marker expression of the MCFC dataset in the monocyte subset. (B) Linear regression between the respective MCFC percentages of CD14+/CD16+ monocytes and age in control and ADD individuals. R (Pearson’s correlation coefficient) and p-values are indicated. (C) Normalized expression of marker genes in the scRNA-seq monocyte space across cell states. (D) Module scores of ADD-derived differentially expressed gene (DEG) signature enrichment per group for the combined *CD14+* space in the scRNA-seq dataset integrated for season of sampling. Statistical analysis using Wilcoxon test on sample level and Benjamini-Hochberg-adjusted p-values are indicated. (E) Linear regression between the median combined module score of DEGs of monocytes and age in control and ADD individuals. R (Pearson’s correlation coefficient) and p-values are indicated. (F) Relative frequencies and absolute cell numbers of states within the monocyte space for the scRNA-seq dataset. Groups were normalized to 1,000 cells. (G) UMAPs of the scRNA-seq monocyte space colored by module scores for the up- or downregulated ADD DEG signatures. Violin plots show module scores per monocyte state and either up or down DEG signature. (H) Radar chart visualization of functional enrichment of DEGs per group compared to controls for the hallmark database term *TNFa signaling via NFkB*. (I) Predicted NF-*k*B pathway activity for all groups using PROGENy in classical monocytes. (J) Expression of the DE genes from the hallmark term *TNFa signaling via NFkB* across different groups. Color indicates scaled average expression, and dot size indicates percent of cells expressing the gene. Genes are ordered by hierarchical clustering.

**Figure S3 –.**
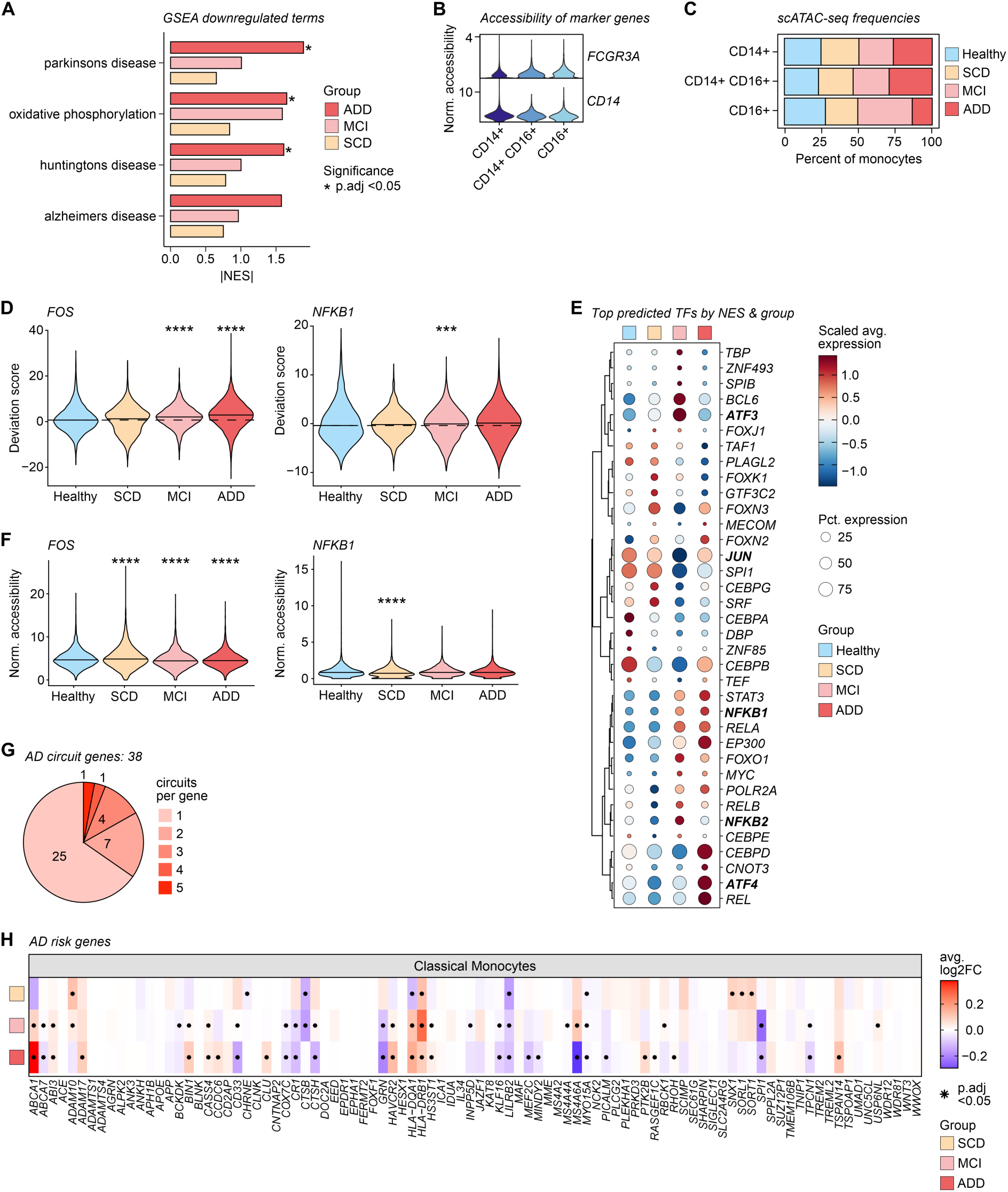
Inflammatory programming of classical monocytes in Alzheimer’s Disease is transcriptionally and epigenetically driven by AP-1 and NF-*k*B. (A) Top downregulated terms from a gene set enrichment analysis (GSEA) per A+ group based on KEGG database terms for classical monocytes. * indicates p-value < 0.05 (Benjamini-Hochberg adjustment). Top 4 terms by |NES| are depicted. NES = normalized enrichment score. (B) Normalized accessibility of the *FCGR3A* (CD16) and *CD14* locus in the scATAC-seq monocyte dataset. (C) Relative frequencies of cell states within the monocyte space for the scATAC-seq dataset. (D) Motif enrichment of selected transcription factors (TFs) identified by scATAC-seq or scRNA-seq in the classical monocyte space. Statistical analysis using Wilcoxon test. *** indicates p < 0.001, **** indicates p < 0.0001. (E) Expression of top TFs from TF binding site prediction with highest normalized enrichment scores (NES) per group. Color indicates scaled average expression and dot size indicates percent of cells expressing the gene. Genes are ordered by hierarchical clustering. (F) Normalized accessibility of the *FOS* and *NFKB1* locus in the scATAC-seq data. Statistical analysis using Wilcoxon test. **** indicates p-value < 0.0001. (G) Distribution of number of regulatory circuits per circuit gene identified by the tool MAGICAL. (H) Heatmap of log2 fold change compared to controls of AD risk genes in classical monocytes across A+ groups (logistic regression framework with correction for season of sampling, * indicates Bonferroni adjusted p-value < 0.05).

**Figure S4 –.**
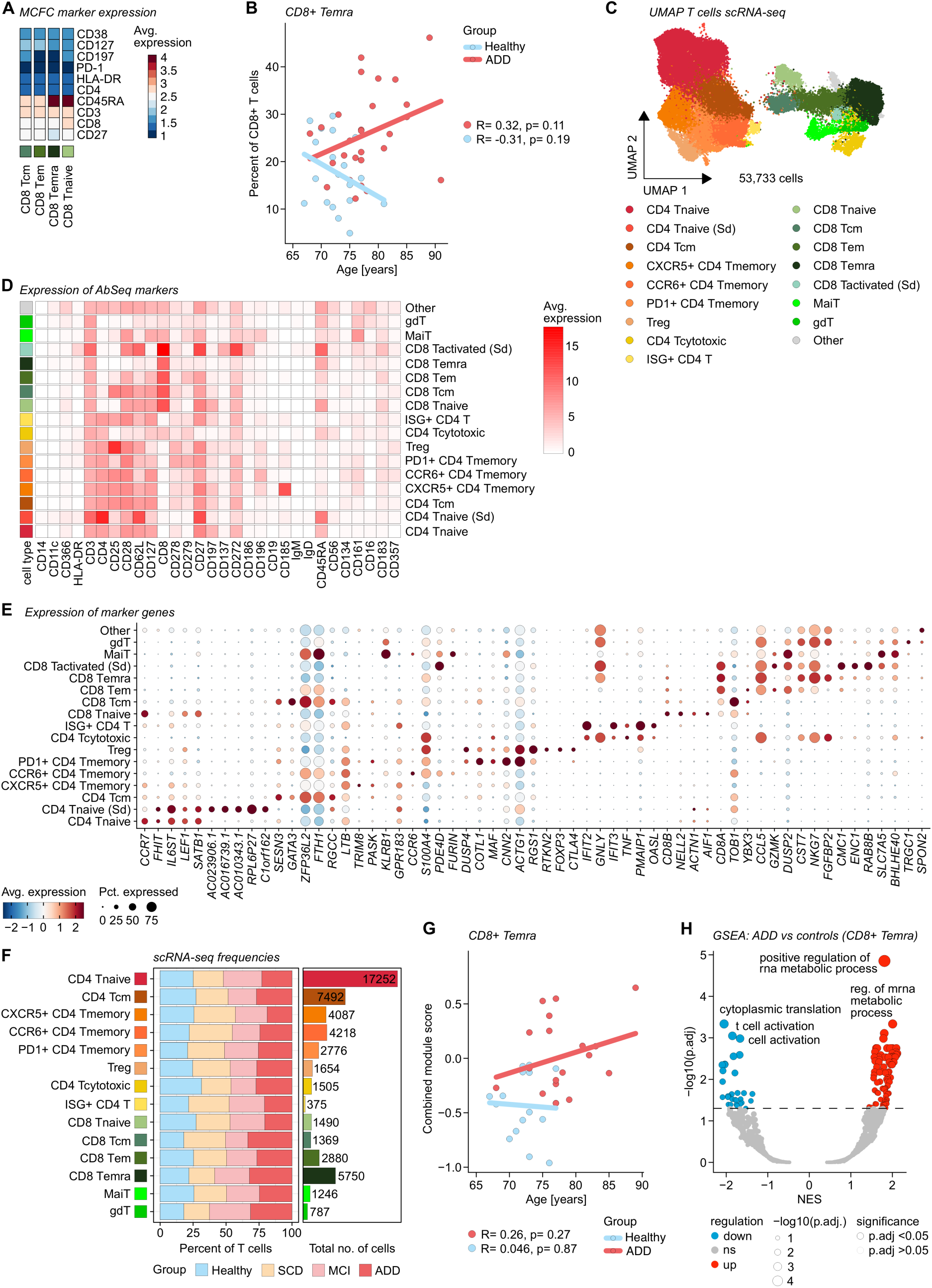
Dysfunctional, terminally differentiated CD8+ T cells expand in Alzheimer’s Disease. (A) Average surface marker expression in the CD8+ compartment of the MCFC dataset. (B) Linear regression between the MCFC percentages of CD8+ Temra cells and age in control and ADD individuals. R (Pearson’s correlation coefficient) and p-values are indicated. (C) Combined UMAP visualization of T cells within the scRNA-seq data colored by cell state (n = 53,733 cells). (D) Average dsb-normalized expression of the AbSeq panel in CD4+ and CD8+ T cell states. (E) Marker gene expression of CD4+ and CD8+ cell state markers within annotated states of the scRNA-seq data. (F) Relative frequencies and absolute cell numbers of CD4+ and CD8+ T cell states within the T cell space for the scRNA-seq dataset. Each group was normalized to 1,000 cells. (G) Linear regression between the median combined module score of DEGs of CD8+ Temra cells and age in control and ADD individuals. R (Pearson’s correlation coefficient) and p-values are indicated. (H) Gene set enrichment analysis (GSEA) on GO biological processes (BP) in CD8+ Temra cells comparing the ADD and control group. NES = normalized enrichment score.

**Figure S5 –.**
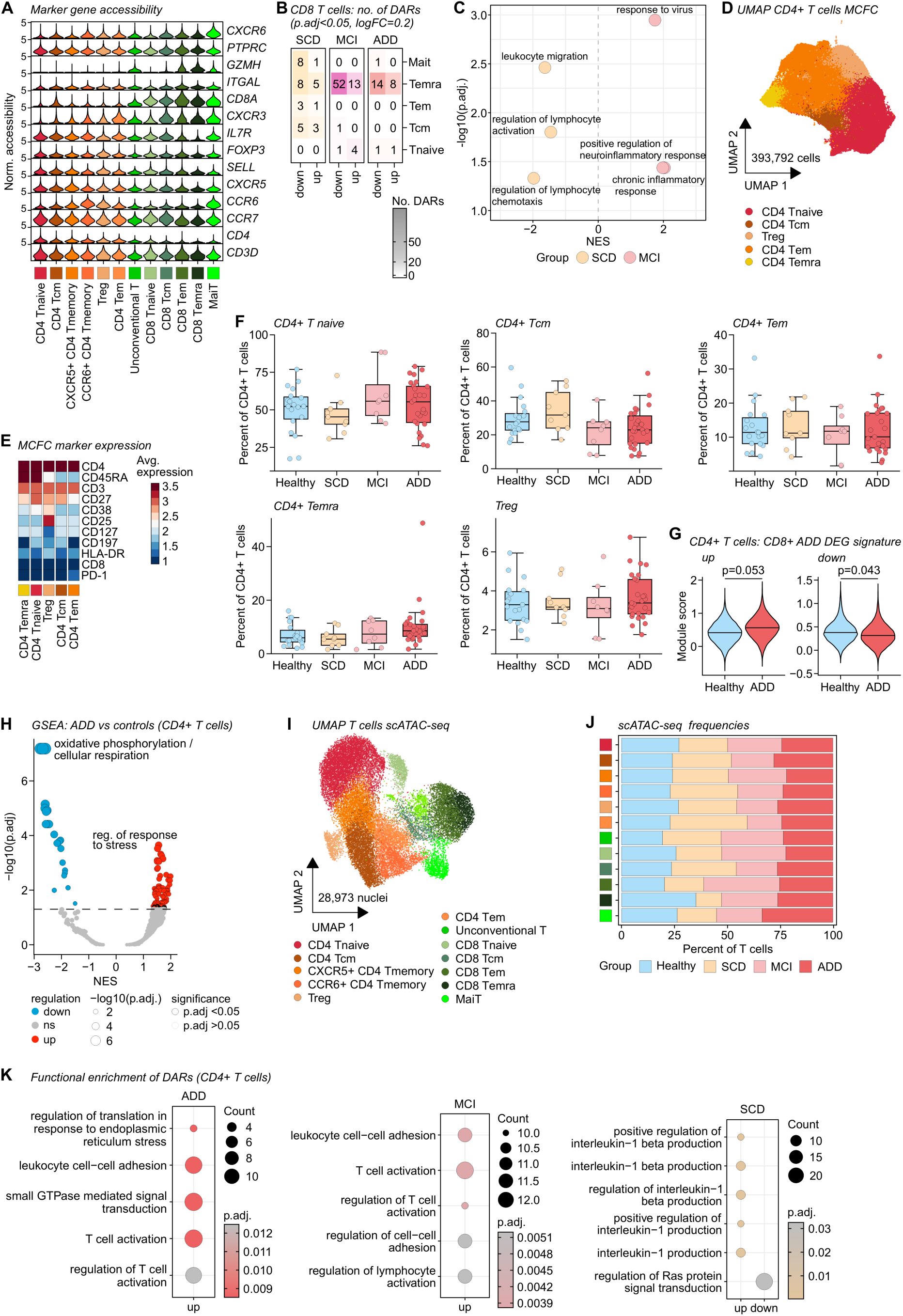
Transcriptional and epigenetic ADD-associated patterns in CD8+ T cells are partially shared in CD4+ T cells. (A) Normalized accessibility of marker gene loci for the CD4+ and CD8+ T cell states annotated in the scATAC-seq dataset. (B) Number of differentially accessible regions (DARs) within CD8+ T cells. (C) Normalized enrichment score (NES) for different GO biological processes (BP) database terms in the chromatin accessibility of CD8+ Temra cells. (D) UMAP visualization of CD4+ T cells in the MCFC dataset colored by cell state (n = 8,614,672 cells were downsampled to 393,792 cells for visualization). (E) Average surface marker expression in the CD4+ compartment of the MCFC dataset. (F) Relative frequencies of CD4+ T cell states as percentage of total CD4+ T cells. Statistical analysis using Kruskal-Wallis and post hoc Dunn’s test. (G) Module score of differentially expressed gene (DEG) signature derived from CD8+ T cells from ADD patients mapped onto CD4+ T cells in the scRNA-seq dataset integrated for season of sampling. Statistical analysis using Wilcoxon test on sample level, p-values are indicated. (H) Gene set enrichment analysis (GSEA) on GO biological processes (BP) in CD4+ T cells comparing the ADD and control group. NES = normalized enrichment score. (I) UMAP visualization of the combined CD4+ and CD8+ T cell compartment in the scATAC-seq dataset (n = 28,973 nuclei). (J) Relative frequencies of CD4+ and CD8+ T cell states within the T cell space for the scATAC-seq dataset. Each group was normalized to 1,000 cells. (K) Functional enrichment of GO biological processes (BP) database terms based on DAR-associated genes in A+ groups in CD4+ T cells.

**Figure S6 –.**
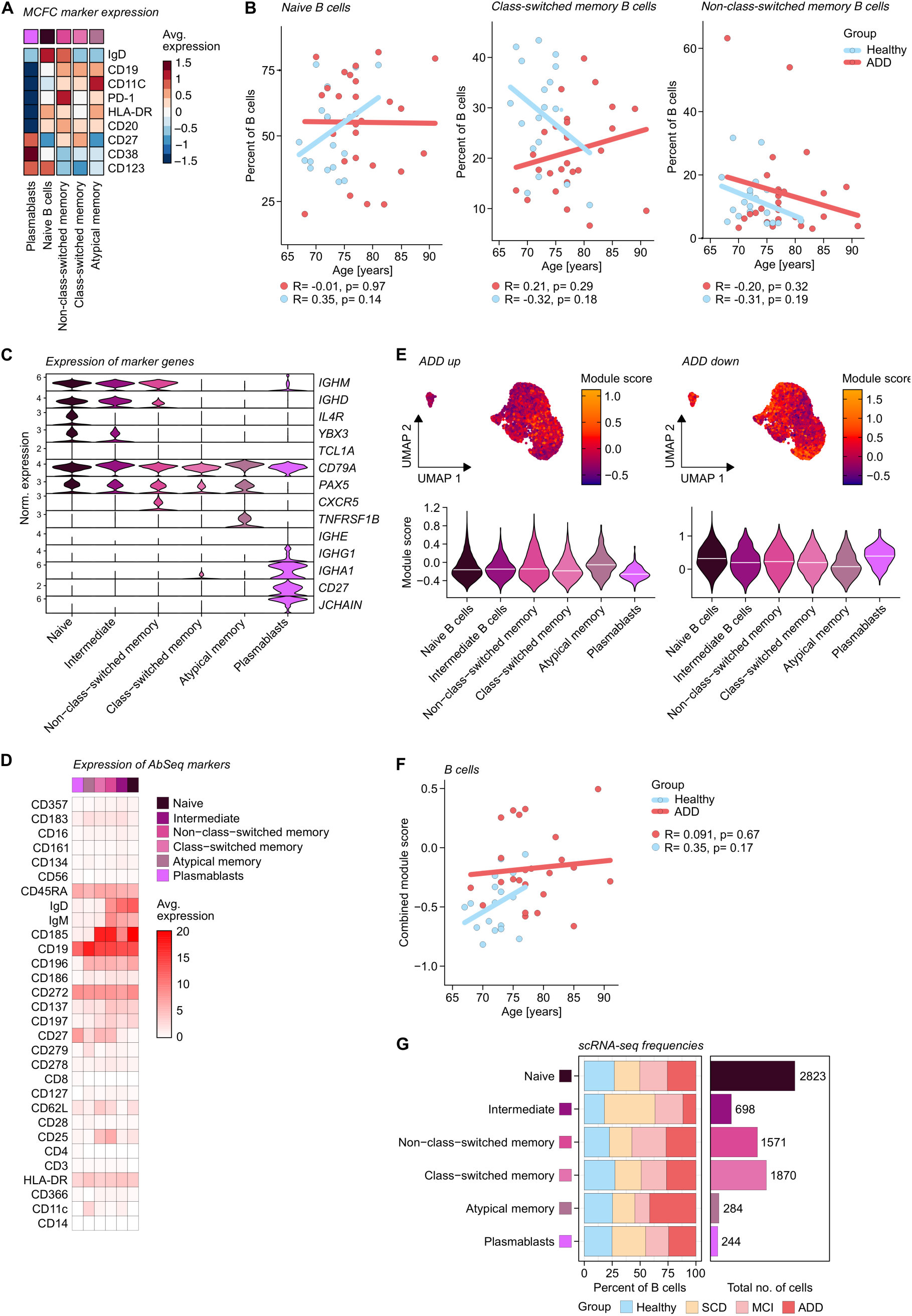
Contraction and loss of function define B cell immunity in Alzheimer’s Disease. (A) Surface marker expression of the different cell subtypes in the MCFC B cell subset. (B) Linear regression between the MCFC percentages of B cell states and age in control and ADD individuals. R- (Pearson’s correlation coefficient) and p-values are indicated. (C) Normalized marker gene expression in the scRNA-seq dataset colored by B cell state. (D) Average dsb-normalized expression of the AbSeq panel in B cell states. (E) UMAPs colored by module score derived from the ADD differentially expressed gene (DEG) signature. Violin plots show module scores per B cell state. (F) Linear regression between the median combined module score of DEGs of B cells and age in control and ADD individuals. R (Pearson’s correlation coefficient) and p-values are indicated. (G) Relative frequencies and absolute cell numbers of the cell states within the B cell space for the scRNA-seq dataset. Groups were normalized to 1,000 cells.

